# Accurately Differentiating COVID-19, Other Viral Infection, and Healthy Individuals Using Multimodal Features via Late Fusion Learning

**DOI:** 10.1101/2020.08.18.20176776

**Authors:** Ming Xu, Liu Ouyang, Yan Gao, Yuanfang Chen, Tingting Yu, Qian Li, Kai Sun, Forrest Sheng Bao, Lida Safarnejad, Jing Wen, Chao Jiang, Tianyang Chen, Lei Han, Hengdong Zhang, Yue Gao, Zhengmin Yu, Xiaowen Liu, Tianyu Yan, Hebi Li, Patrick Robinson, Baoli Zhu, Jie Liu, Yang Liu, Zengli Zhang, Yaorong Ge, Shi Chen

## Abstract

Effectively identifying COVID-19 patients using non-PCR clinical data is critical for the optimal clinical outcomes. Currently, there is a lack of comprehensive understanding of various biomedical features and appropriate technical approaches to accurately detecting COVID-19 patients. In this study, we recruited 214 confirmed COVID-19 patients in non-severe (NS) and 148 in severe (S) clinical type, 198 non-infected healthy (H) participants and 129 non-COVID viral pneumonia (V) patients. The participants’ clinical information (23 features), lab testing results (10 features), and thoracic CT scans upon admission were acquired as three input feature modalities. To enable late fusion of multimodality data, we developed a deep learning model to extract a 10-feature high-level representation of the CT scans. Exploratory analyses showed substantial differences of all features among the four classes. Three machine learning models (k-nearest neighbor kNN, random forest RF, and support vector machine SVM) were developed based on the 43 features combined from all three modalities to differentiate four classes (NS, S, V, and H) at once. All three models had high accuracy to differentiate the overall four classes (95.4%-97.7%) and each individual class (90.6%-99.9%). Multimodal features provided substantial performance gain from using any single feature modality. Compared to existing binary classification benchmarks often focusing on single feature modality, this study provided a novel and effective breakthrough for clinical applications. Findings and the analytical workflow can be used as clinical decision support for current COVID-19 and other clinical applications with high-dimensional multimodal biomedical features.

**One sentence summary:** We trained and validated late fusion deep learning-machine learning models to predict non-severe COVID-19, severe COVID-19, non-COVID viral infection, and healthy classes from clinical, lab testing, and CT scan features extracted from convolutional neural network and achieved predictive accuracy of > 96% to differentiate all four classes at once based on a large dataset of 689 participants.

## Introduction

The COVID-19 is an emerging and major biomedical challenge to the entire healthcare system (WHO 2020). Currently, the confirmatory diagnosis of COVID-19 is based on the qRTPCR and/or antigen testing for the SARS-CoV-2 virus (NHC 2020, Xiao et al. 2020, Wang et al. 2020). While these tests are the gold standard of COVID-19 diagnosis, they suffer from reliability, resource adequacy, reporting lag, and testing capacity issues (Boger et al. 2020). To help frontline clinicians diagnose COVID-19 more effectively and efficiently, other diagnostic methods have also been explored and utilized, including medical imaging (e.g., X-ray and CT scans; Zhou et al. 2020), lab testing results (e.g., various blood biochemistry; Fu et al. 2020, Shi et al. 2020, Ojha et al. 2020, Lyu et al. 2020), and more commonly used clinical symptoms (Brooks 2020). These methods do not directly detect the disease-causing SARS-CoV-2 virus (or its antigen); therefore, they do not have as conclusive power as confirmatory molecular diagnostic methods. Nevertheless, these alternative methods help clinicians with inadequate resources and capability to detect COVID-19, differentiate COVID-19 patients from other non-COVID patients, and triage to optimize the resources in healthcare facilities (Wu and McGoogan 2020, Troug et al. 2020).

Currently, analyzing CT scans can differentiate COVID-19 patients, especially those in severe clinical state, from healthy people or patients with non-COVID infections. COVID-19 patients usually present typical ground glass opacity (GGO) characteristic on their CT scan image. One study reported a 98% detection rate of COVID-19 based on a 51-patient sample and without a comparison group (Fang et al. 2020). Detection rates between 60%-93% were observed in another study based on 1014 patients with a comparison group (Ai et al. 2020). The recent advances in data-driven deep learning methods, such as convolutional neural networks (CNNs), have been demonstrated to detect COVID-19 patients. However, the effectiveness of using deep learning methods to further differentiate COVID-19 and other clinically similar non-COVID viral infections still needs to be explored. Certain regions of the world, including Hubei, China in February 2020, adopted CT scan as the official clinical diagnosis in addition to the molecular confirmatory diagnosis of COVID-19, according to the nation’s diagnosis and treatment guidance (NHC 2020).

Similarly, for places where molecular confirmatory diagnoses are not immediately available, symptoms are often used for quickly evaluating presumed patients’ condition and helping triage (Menni et al. 2020, Timmers et al. 2020, Troug et al. 2020). There are checklists developed for self-evaluating the risk of developing COVID-19 based on clinical information including symptoms, pre-existing comorbidities, and various demographic, behavioral, and epidemiological factors. However, these clinical data are generally used for qualitative purposes (e.g., initial assessment) for both the general public and clinicians (Nair et al. 2020). Their effectiveness as accurate diagnostic decision support is largely underexplored and remains unknown.

In addition to biomedical imaging and clinical information, more recent studies on COVID-19 have shown that laboratory testing results such as various blood biochemistry are also a feasible candidate to detect COVID-19 patients with reasonably high accuracy (Brinati et al. 2020, Sun et al. 2020). The rationale is that the human body is a unity. When infected with COVID-19, the clinical consequence can be shown not only from apparent symptoms but also hematological biochemistry change. Because of the challenge of asymptomatic infection of COVID-19, other types of biomedical information such as lab testing results can be alternative diagnostic decision support evidence. It is possible that our current definition and/or understanding of “asymptomatic infection” may be extended by more intrinsic, quantitative, and subtle biomedical evidence (Daniells et al. 2020, Gandhi et al. 2020).

Despite tremendous advances of alternative and complementary diagnostic evidence for COVID-19, there are still substantial clinical knowledge gaps and technical challenges that hinder our efforts on harnessing the power of various biomedical data. First of all, most current studies usually focus on one type (modality) of multiple modalities of diagnostic data and do not consider the potential interactions and added interpretability among them. For example, can we leverage both CT scan and clinical information to develop a more accurate COVID-19 diagnostic decision support system (Shi et al. 2020)? As stated earlier, the human body is a unity against SARS-CoV-2 infection. Biomedical imaging and clinical approaches evaluate different aspects of the clinical consequences of COVID-19. By combining these different modalities of biomedical information, we may be able to achieve a more comprehensive characterization of COVID-19. This is referred to as a “*multimodal* biomedical information” research.

Secondly, while there are ample accurate deep learning (DL) algorithms/models/tools especially using biomedical imaging, most of them focus on the efforts of differentiating COVID-19 from non-infected healthy individuals. A moderately trained radiologist can differentiate CT scans of COVID-19 patients from healthy individuals with high accuracy as well, making the current efforts of developing DL algorithms not clinically useful for the binary classification problem (Fang et al. 2020). The more critical and urgent clinical question is not only to differentiate COVID-19 from non-infected healthy individuals but also from other non-COVID viral infections (Li et al. 2020, Altmayer et al. 2020). Patients with non-COVID viral infection also present GGO in their CT scans. Therefore, the specificity of GGO as diagnostic criteria of COVID-19 is low (Ai et al. 2020). In addition, both non-severe COVID-19 and non-COVID viral infection patients share some easily confusing common symptoms (Qu et al. 2020). For frontline clinicians, effectively differentiating non-severe COVID-19 from non-COVID viral infection is therefore a challenging task without confirmatory molecular testings, which may not be readily available at the time of admission. Similarly, differentiating asymptomatic and pre-symptomatic (including those in non-severe clinical type) COVID-19 from non-infected healthy individuals is another major clinical challenge (Ooi and Low 2020). There is a more urgent need for a *multinomial* classification system that can detect COVID-19 (including those asymptomatic), non-COVID viral infection, and healthy individuals all at once, rather than running several binary classifiers in parallel independently (Baltruschat et al. 2019).

The third major challenge comes from a more computational aspect. We should harness the power of multimodal biomedical information from imaging, clinical information, and lab testing data altogether. In theory, it will provide a more comprehensive understanding of COVID-19, which leads to a more accurate decision support system that can differentiate multinomial COVID-19 (severe and non-severe), non-COVID viral infection, and healthy individuals all at once. However, biomedical imaging CT data with high-dimensional feature space do not integrate well with low-dimensional clinical and lab testing data. Current studies usually only describe the association between biomedical imaging and clinical features (Ai et al. 2020, Song et al. 2020, Wu et al. 2020, Meng et al. 2020, Cheng et al. 2020, Zhu et al. 2020), and the potential power for an accurate decision support tool has not been reported. Technically, CT scan is usually processed with deep learning (DL) methods including CNNs, independent of other types of biomedical data. Low-dimensional clinical and lab testing data are usually analyzed with traditional hypothesis-driven methods (e.g., binary logistic regression or multinomial classification) or other non-DL machine learning methods such as random forest (RF), support vector machine (SVM), and k-nearest neighbor (kNN). The huge discrepancy of feature space dimensionality between CT scan and clinical/lab testing data makes it an especially challenging task of multimodality fusion to combine these different aspects of biomedical information directly (Balrtusaitis et al. 2019).

To fill these knowledge gaps and overcome the technical challenge of analyzing multimodal biomedical information effectively, we propose the following study objective.

Clinically, we aim to accurately differentiate among non-severe COVID-19, severe COVID-19, non-COVID viral pneumonia, and healthy individuals all at once.

To successfully fulfill this much demanded clinical objective, we will develop a novel hybrid deep learning-machine learning framework via feature late fusion that harnesses the power of a wide array of complex multimodal data. The clinical objective and technical approach in this study synergistically complement each other to form the basis of an accurate COVID-19 diagnostic decision support system.

## Method

### Participant Recruitment

We recruited a total of 362 confirmed COVID-19 patients with two independent qRTPCR tests from Wuhan Union Hospital between January 2020 and March 2020 in Wuhan, Hubei Province, China. For this study we did not aggregate COVID-19 patients under the same class because the clinical characteristics and potential prognosis (e.g., death) were distinct between non-severe and severe states. The recruited COVID-19 patients were further categorized as 148 severe and 214 non-severe clinical states. Patients in the severe state group were identified by having one of three clinical features: 1) respiratory rate > 30 breaths per minute; 2) oxygen saturation < 93% at rest; and 3) arterial oxygen partial pressure (PaO2)/fraction of inspired oxygen (FiO2) < 300mmHg (40kPa), according to the official COVID-19 Diagnosis and Treatment Plan from the National Health Commission of China (NHC 2020) as well as guidelines from American Thoracic Society (Metlay et al. 2019). The non-infected group included 198 healthy individuals without any infections. These participants were the 2019 Hubei Provincial CDC regular annual physical examination cohort. This group represented a baseline healthy group and was mainly used to compare and contrast with non-severe COVID-19 patients, especially those showing inconspicuous clinical symptoms.

In order to differentiate COVID-19 especially non-severe COVID-19 from other clinically similar non-COVID viral infection patients, we also included another group of 129 patients diagnosed with non-COVID viral pneumonia. Note that the term “viral pneumonia” was an umbrella term including diseases caused by more than one type of virus, such as influenza and adenovirus. However, in clinical practice for this study, it would be adequate to detect and differentiate COVID-19 from non-COVID viral infections for initial triaging. We recruited 129 confirmed non-COVID viral infection participants from Kunshan Hospital, Suzhou, China. The reality was that most healthcare resources were optimized for COVID-19 and some patients presenting COVID-like symptoms or having GGOs were clinically diagnosed as COVID-19 without confirmatory qRT-PCR test in Hubei, especially during February 2020. Therefore, it was not possible to recruit non-COVID viral infection participants in Hubei during the similar period as we recruited COVID-19 patients.

In summary, the entire study participants comprised four mutually exclusive multinomial classes: severe COVID-19 (S) with 148 participants, non-severe COVID-19 (NS) with 214 participants, non-COVID viral infection (V) with 129 participants, and non-infected healthy (H) with 198 participants. This study was in full compliance with the Declaration of Helsinki. It was rigorously evaluated and approved by both IRB committees of Wuhan Union Hospital, Huazhong University of Science and Technology (approval number 2020-IEC-J-345) and Kunshan People Hospital, Jiangsu Provincial Center for Disease Control and Prevention (approval number JSJK2020–8003–01). All participants were comprehensively explained about the study and signed a written informed consent before being admitted.

### Medical Feature Selection and Description

Patient participants (including S, NS, and V classes) were screened upon initial admission into hospitals. Their clinical information, including pre-existing comorbidity, symptoms, demographic, epidemiological and other clinical data were recorded. For the non-infected healthy class (H), participants’ clinical data were extracted from the Hubei Provincial CDC’s physical examination record system. Patient-level sensitive information, including name and exact residency were completely deidentified. After comparing among these different classes, the following 23 clinical features were selected: smoking history (SMK), hypertension (HYP), type-2 diabetes (DIA), cardiovascular disease (any type, CAR), COPD (CPD), fever (FEV), low fever (LOF), medium fever (MDF), high fever (HIF), sore throat (SOR), coughing (COU), phlegm production (MUC), headache (HED), feeling chill (CHL), muscle ache (MSA), feeling fatigue (FTG), chest congestion (SHB), diarrhea (DIR), loss of appetite (NAP), vomiting (VOM), elderly (>50yr dichotomized and encoded as old, OLD), and gender (SEX). These clinical data were dichotomized as either having the condition (1) or not having the condition (0; Fig. 1). Note that some clinical features, especially symptoms, were self-reported by the patients. A more comprehensive definition and description of clinical features were provided in supplementary table SI1. Prevalence (number of participants having a given feature over total participants in the class) of each clinical feature were computed across four classes. For the 0–1 binary clinical features, pairwise *z*-test was then applied to detect any substantial between-class difference of prevalence (i.e., proportion) of these features.

**Fig. 1.**
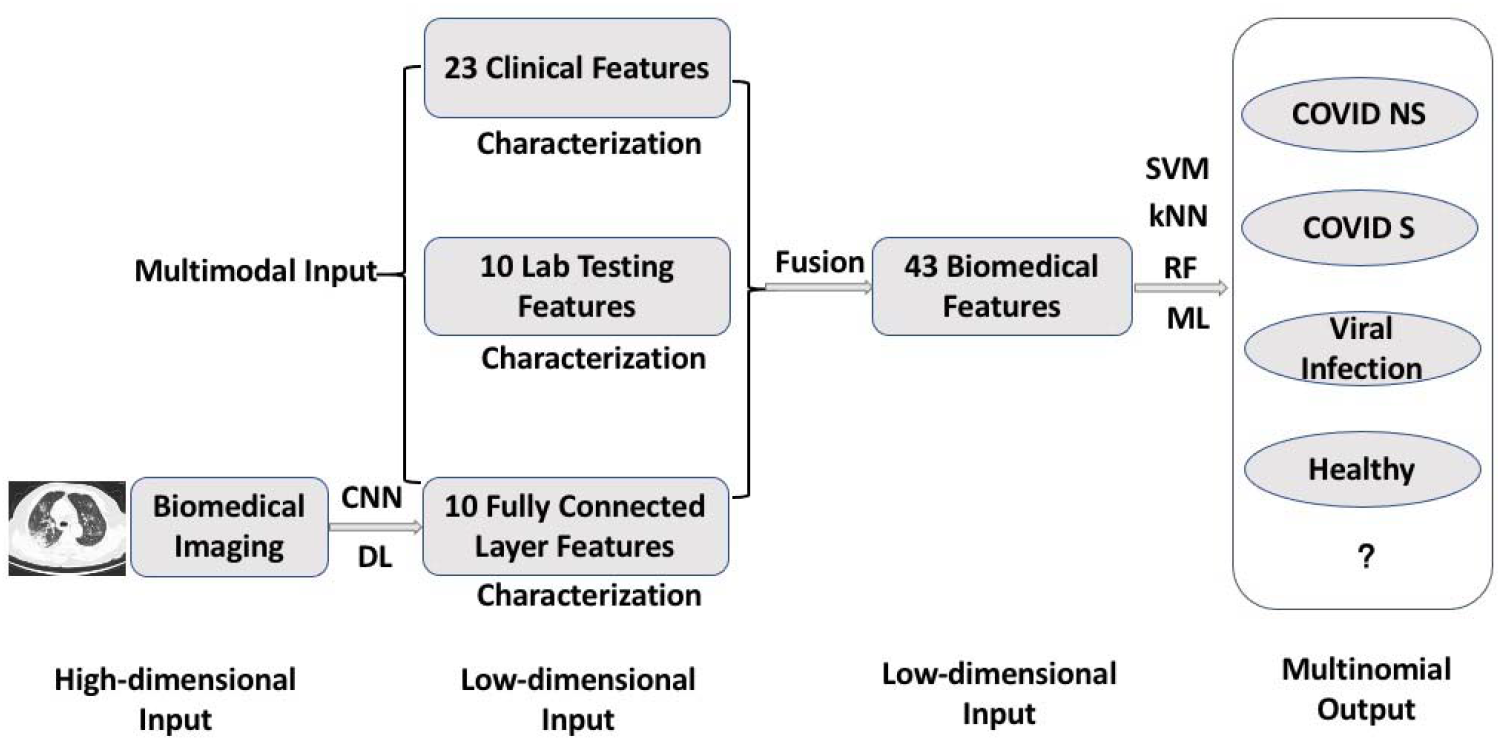
Multimodal Feature Late Fusion and Multinomial Classification Workflow.

Note: deep learning (DL) convolutional neural network (CNN) was performed on CT scan images for representation learning and extracting 10 features from a customized fully connected layer. These 10 features were merged with other modality data and known as feature late fusion. In the machine learning (ML) stage of the workflow, each of the three ML models (support vector machine SVM; k-nearest neighbor kNN; and random forest RF) worked independently to provide their respective outputs.

The lab testing features were extracted from participants’ electronic health record (Fig. 1). Only lab tests done at the time of admission were included. Non-infected healthy participants’ blood samples were taken during their annual physical examination. We selected lab testing features that were present in at least 90% of participants in any of the four classes (severe COVID-19, non-severe COVID-19, non-COVID viral infection, and non-infected healthy participants). After screening, the following 10 features were included: white blood cell (WBC), hemoglobin (HGB), palette (PLT), neutrophil (NE), neutrophil percent (NE%), lymphocyte (LY), lymphocyte percent (LY%), C-reactive protein (CRP), total bilirubin (TBIL), and creatine (CREA). Features in the lab testing modality all had continuous numeric values, different from 0–1 binary values in the clinical feature modality. Distributions of these lab testing features were compared across the four classes using two-sided Kolmogrov-Smirnov test. In addition, we also applied Kruskal-Wallis test for multiple comparisons across the four classes for the top three most differentiating features identified by later machine learning workflow. Kolmogrov-Smirnov test was applied for initial screening to investigate whether the value of the same biomedical feature distributed differently between two classes. Non-parametric Kruskal-Wallis test was chosen for more rigorous between-class comparisons because the Kruskal-Wallis test was more robust against non-normality in the data and was able to accommodate more than two classes, i.e., multinomial, in this study.

Each participant took CT scans in the thoracic region in the radiology department. The CT scan imaging was archived and presented in the DICOM (Digital Imaging and Communications in Medicine) format. Toshiba Activion 16 multislice CT scanners were used to perform CT scanning at around 120kVp and 50mA tube current. There were 50 CT images per scan, slice thickness was 2mm, voxel size 1.5mm, and image resolution was 512×512 pixel.

### The Multinomial Classification Objective

The main research goal in this study is to accurately differentiate COVID-19 in both severe and non-severe clinical states, non-COVID viral infection, and non-infected individuals from a total of *N* participants all at once. Therefore, it was formulated as a *multinomial* output classification problem with one of the four mutually exclusive output classes H (non-infected healthy); V (non-COVID viral pneumonia); NS (non-severe COVID-19); and S (severe COVID-19) of an individual *i*:

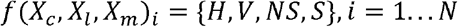

where the inputs were individual *i*’s *multimodal* features of binary clinical information (*Xc*), continuous lab testing results(*Xl*), and CT scan imaging (*Xm*).

The major advantage of this study was to classify four classes all at once instead of developing several binary classifiers in parallel.

### The Hybrid Machine Learning-Deep Learning Approach: Feature Late Fusion

As stated earlier, the voxel level CT imaging data does not integrate well with low-dimensional clinical and lab testing features. In this study, we propose a feature late fusion approach via a hybrid of deep learning and machine learning models. Technically, deep learning (DL) is a type of machine learning (ML) which uses deep neural networks (DNNs; CNN was a type of DNN). We colloquially used the term ML as more traditional, non-DL types of ML (e.g., random forest) in this study as a contrast to DL that focused on DNNs. An important consideration to successful late fusion of multimodality features was representation learning of the high-dimensional CT features.

First, for CT scan of each participant, we constructed a customized ResNet (Ardakani et al. 2020, Hu et al. 2020, Ko et al. 2020, Li et al. 2020), a specific architecture of DL convolutional neural networks (CNN). By constructing ResNet we were able to transform the voxel level imaging data into a high level representation with significantly fewer features. After several convolution and max pooling layers, ResNet reached a fully connected (FC1) layer before the final output layer. In the commonly used ResNet architecture, the FC layer was a 1×512 vector, which was closer in dimensionality as clinical information (1×23) and lab testing (1×10) feature modalities. However, the original FC layer from ResNet CNN was still much larger than the other two modalities. Therefore, we added another FC layer (FC2) after FC1 but before the final output layer. In this study, FC2 was set to be in 1×10 dimension, i.e., having 10 elements in the vector. Computationally, FC2 served as the low-dimensional, high level representation of the original CT scan data. Distributions of the 10 features in FC2 were compared across the four classes with Kolmogrov-Smirnov test. The technical details of ResNet architecture were provided in supplementary Fig. S1.

Once low-dimensional high-level features were extracted from CT scan data via ResNet CNN, we performed the multimodal feature fusion. For each participant *i*, the clinical information, lab testing, and FC2 features were combined as a single 1×43 (1x[23+10+10]) row vector. The true values of the output were the true observed classes of the participants. Technically, the model would try to predict the outcome as close as the observed classes.

### The Hybrid Machine Learning-Deep Learning Approach: Modeling

After deriving the feature matrix, we then applied ML models for this multinomial classification task. Three different types of commonly used ML models were considered in this study: random forest (RF), support vector machine (SVM), and k-nearest neighbor (kNN). RF is a decision-tree based ML model and the number of tree hyperparameter was set at 10, a relatively small number compared to the number of input features to avoid potential model overfitting. Other RF hyperparameters in this study included Gini impurity score to determine tree split, at least 2 samples to split an internal tree, and at least 1 sample at a leaf node (all default hyperparameter settings from scikit-learn library in Python; default hyperparameters for SVM and RF as well). SVM is a model of maximum hyperplane and L-2 penalty, radial basis function (RBF) kernel, and gamma = 1/43 (inverse of total number of features) were used as hyperparameter values in this study. kNN is a non-parametric instance-based model and k = 5, uniform weights, tree leaf size = 30, p = 2 hyperparameter values were used in this study. These three are technically distinct types of ML models. We aim to investigate whether specific types of ML models and/or multimodal feature fusion would contribute to an accurate COVID-19 classifier for clinical decision support.

We evaluated each respective ML model on 100 independent runs, each run with a different randomly selected set of 80% data for training and the remaining 20% for testing and validation of the model. Having multiple runs instead of one single run was able to reveal model robustness against system stochasticity. The 80%-20% split of the original data to separate training and testing sets also ensured against potential model overfitting and increased model generalizability. In addition, RF models used bagging for internal validation via out-of-bag errors (i.e., how the “tree” would split out in the “forest” model).

After each run, important ML performance metrics, including accuracy, sensitivity, precision, and F1 score were computed for the test set. We reported the overall performance of the ML models first. These different metrics evaluated ML models from different aspects. In this study, we also considered three different approaches to calculate the overall performance for multinomial outputs: micro (one-vs-all), macro (unweighted average, i.e., giving each of the four classes the same 25% weights), and weighted average based on the percentage of each class in the entire sample.

In addition, because the output in this study was multinomial instead of binary, each class would further have its own performance metrics. Then we aggregated the performance metrics across 100 independent runs, determined each metric’s distribution, and evaluated model robustness from these distributions. In general, if ML performance metrics on the testing set had a small variation (i.e., small standard errors), then the model was robust against model input changes and revealed the intrinsic pattern of the data. This was because in each run, a different randomly selected 80% data were selected to train the model.

An advantage of RF over SVM and kNN was that it had relatively clearer interpretability. After developing the RF model, we were able to rank input feature’s importance based on their respective Gini impurity score from the RF model (Chen et al. 2020, Elaziz et al. 2020). We would then explore top contributing features’ clinical relevance to COVID-19.

Finally, we also developed and evaluated the performance of single modality (using clinical information, lab testing, and CT features individually) ML models as baseline conditions. Their performances were then compared with the multimodal classification to demonstrate potential performance gain of feature fusion from different feature modalities.

The deep learning CNN and late fusion machine learning codes were developed in Python with various supporting packages such as scikit-learn. All codes and de-identified data files were freely available on GitHub (https://github.com/forrestbao/corona/tree/master/ct).

## Results

### Clinical Differences among Four Classes

We compared clinical features across four classes. The prevalences of each feature in all four classes were shown in supplementary Fig. S2. In general, most clinical features varied substantially among non-severe COVID-19 (NS), severe COVID-19 (S), and non-COVID viral pneumonia (V) classes. Note that all symptom features in the healthy (H) class were zero as reference, except gender and age group (>50yr). Based on two-sample z-test of proportions, NS and S classes differed significantly (*p*< 0.05) in 10 out of 22 symptom features, most of which were comorbidities such as hypertension, diabetes, cardiovascular diseases, and COPD. Comparing between NS and V classes, they differed significantly in 12 features, including smoking habitat, fever, sore throat, etc. However, NS and V classes did not differ significantly in comorbidities. S and V differed significantly in 16 out of 22 features, making these two classes the most distinct based on symptoms. These results showed that the prevalences of clinical features differed substantially among the classes. Complete z-test results of each clinical feature between each pair of classes were provided in supplementary Table S1.

In addition, we also showed the top three differentiating clinical features based on later machine learning random forest (RF) analysis: fever, coughing, and elderly (>50yr). For fever and coughing, we used V class as the reference and constructed 2×2 contingency tables between NS and V, as well as S and V. We demonstrated the odds ratio (OR) and 95% confidence interval (95% CI) on the forest plot in Fig. 2 (left panel). Compared to patients in V class, patients in both NS and S class were more likely to develop fever (>37C). In addition, S patients also experienced more fever than NS based on the forest plot. Therefore, fever was one of the major determining factors to differentiate multiple classes. Next, both NS and S patients reported significantly less coughing than in V patients (Fig. 2, middle panel). There were no statistically significant differences between NS and S. Finally, for the elderly feature, we included all three S, NS, and H classes because the elderly prevalence of H class was not zero. The forest plot was shown in Fig. 2 (right panel). S patients were significantly older than V patients while NS and H were younger than V patients. These differences in clinical features among the four classes would pave the way towards a data-driven machine learning model.

**Fig. 2.**
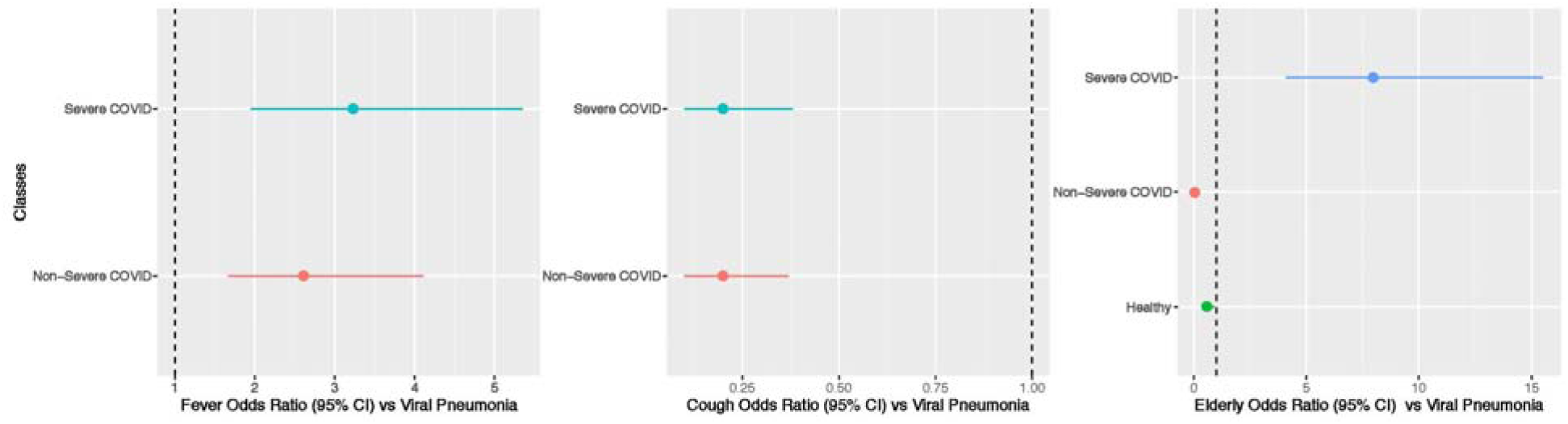
Forest Plot of Top Three Differentiating Clinical Features.

Note: Viral pneumonia (V) was the reference class to compare with and to calculate odds ratios (ORs). Healthy class had no individuals showing fever or cough, hence were not included in the first two panels (left and middle). The error bars represented variation in estimating ORs, not the original feature variations.

### Lab Testing Differences among Four Classes

For the continuous lab testing features, we calculated and compared the exact distributions among four classes. The boxplot of each lab testing feature across four classes were provided in supplementary Fig. S3. In general, the four classes differed substantially across many lab testing features. Based on the two-sided Kolmogov-Smirnov test (KS-test) results, NS and S classes were only similar on hemoglobin and palettes and differed significantly in all remaining eight features. In other words, NS and S patients’ lab testing features had distinct distributions. Similarly, NS and H were only similar with regard to creatine; NS and V were only similar in hemoglobin, palettes, and total bilirubin; S and H were only similar in total bilirubin; S and V were only similar in hemoglobin and neutrophils; V and H were only similar in white blood cell counts. The complete KS-test results of each lab testing feature between each pair of classes were provided in supplementary Table S1.

The three most influential differentiating features based on RF modeling were CRP, hemoglobin, and neutrophil. The distributions of CRP among four classes were provided in the boxplot in Fig. 3 (left panel). In addition to the KS-test that did not account for multiple comparison across classes, further pairwise comparisons were further performed by the nonparametric Kruskal-Wallis (KW) H test. Each of the six pairs as well as the overall KW tests showed significant between-class differences. The distributions of hemoglobin (HGB) were shown in Fig. 3 (middle panel). While H differed significantly from NS, S, and V with regard to HGB, the other three pairs did not show statistically significant differences. The distributions of neutrophil (NE) were shown in Fig. 3 (right panel). All pairwise comparisons and the overall KW test showed significant differences between classes.

**Fig. 3.**
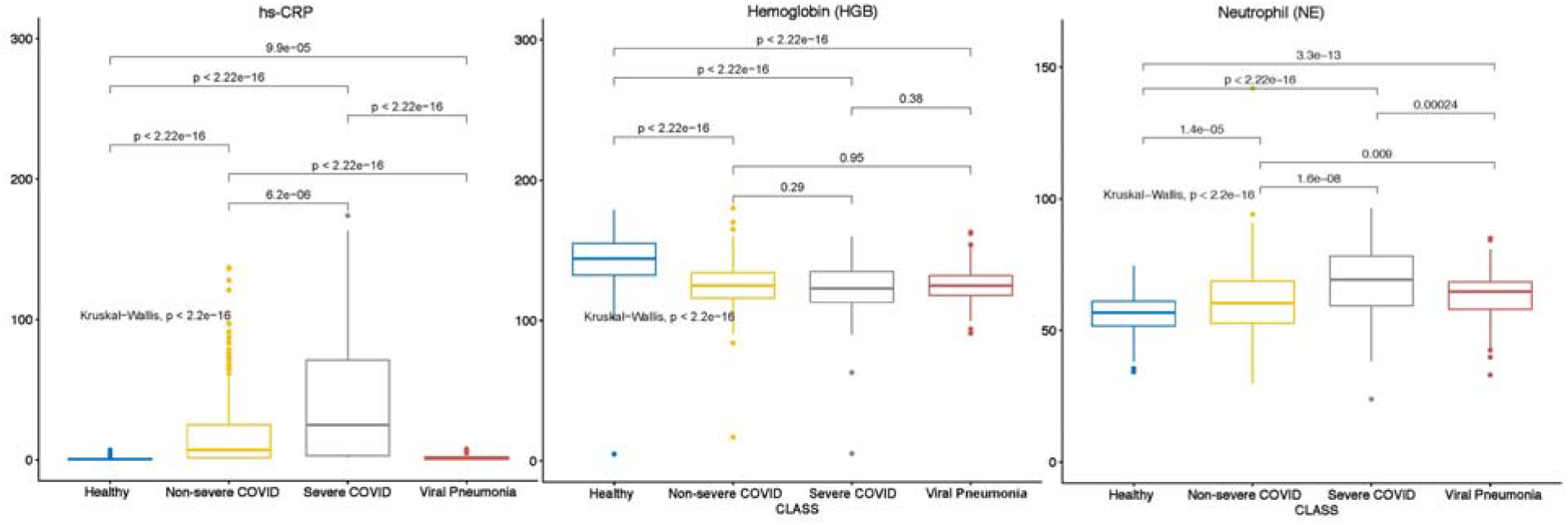
Multiple Comparison of Top Differentiating Lab Testing Features.

### CT Differences among Four Classes Based on High Level CNN Features

We analyzed the fully connected layer 2 (FC2) features from ResNet CNN in relation to four classes. The boxplot was shown in supplementary Fig. S4. Two-sided KS-tests showed significant differences between every pair of classes in almost all 10 features in FC2. The only exceptions were feature 6 (CNN6) between S and V class, feature 1, 4, and 5 between H and V classes (supplementary table S2). Based on the RF results, feature 1, 6, and 10 were the three most critical features in FC2 for multinomial classification. Further KW tests were performed on these three features and the results were shown in Fig. 4. These results demonstrated the possibility of an accurate classifier based on CNN representation of high level features.

**Fig. 4.**
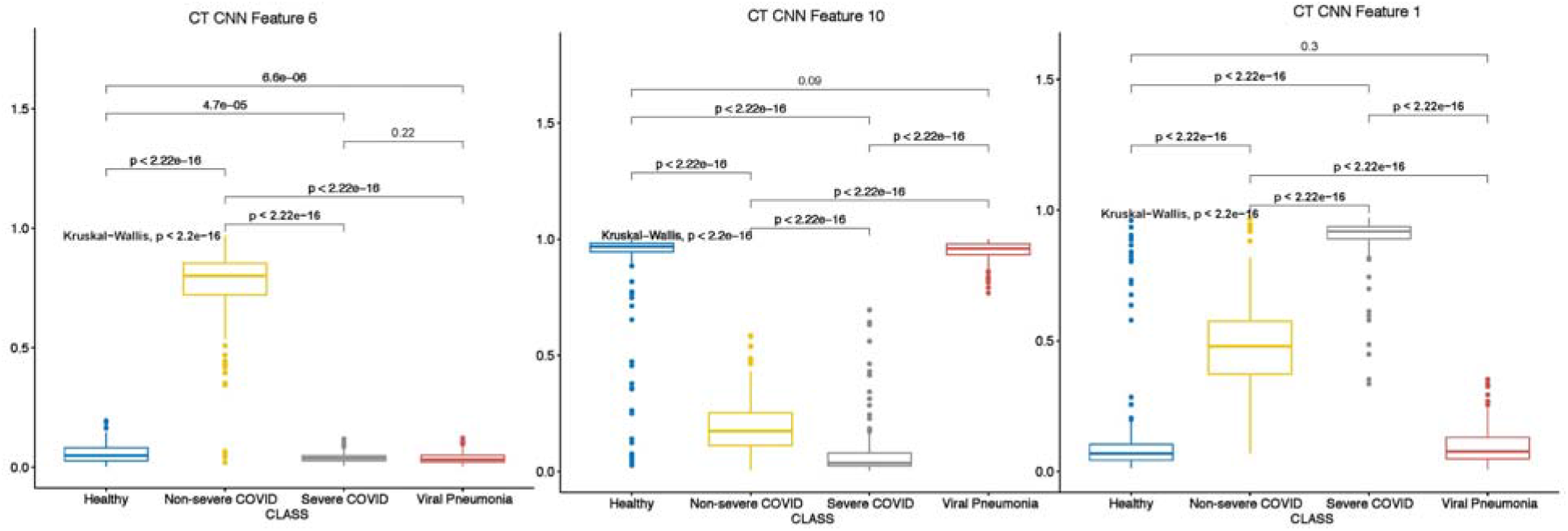
Multiple Comparison of Top Differentiating CT Features via CNN.

### Accurate Multimodal Model for COVID-19 Multinomial Classification

We developed and validated three different types of ML models: kNN, RF, and SVM. Overall multimodal classification accuracy was 96.2% (standard error: ±0.5%), 99.8% (±0.3%),and 99.2% (±0.2%) on average on training data and 95.4% (±0.2%), 96.9% (±0.2%), and 97.7% (±0.1%) on test data, respectively for these three ML models (kNN, RF, and SVM; see Fig. 5). These three models also achieved consistent and high performance based on different approaches of calculating the overall performance, including micro (one-vs-all), macro (unweighted average across all four classes), and weighted average (based on percent of each class) across all four classes. Note that overall accuracy did not depend on sample size so there was only one approach of calculating accuracy. F1 score, sensitivity, and precision each had three different approaches (micro, macro, and weighted average) to quantify. F1 score using macro approach was 95.9% (±0.1%), 98.8% (<±0.1%), 99.1% (<±0.1%) for kNN, RF, and SVM models, respectively. F1 score using micro approach was 96.2% (<±0.1%), 98.8% (<±0.1%), 99.2% (<±0.1%) for kNN, RF, and SVM models, respectively. F1 score using the weighted average approach was 96.2% (<±0.1%), 98.9% (<±0.1%), 99.2% (<±0.1%) for kNN, RF, and SVM models, respectively. The differences of F1 scores based on different approaches (micro, macro, and weighted) were minimal (Fig. 5, comparing across upper, middle, and bottom panels). In addition, the differences of F1 score across different ML models (Fig. 5, represented by different greyscales for a metric) were also insignificant. Similarly, model sensitivity and precision were all > 95% for any ML model type and any approach to calculating the performance metric. Complete overall performance metrics across three different evaluation approaches and three ML models were recorded in supplementary table S3.

**Fig. 5.**
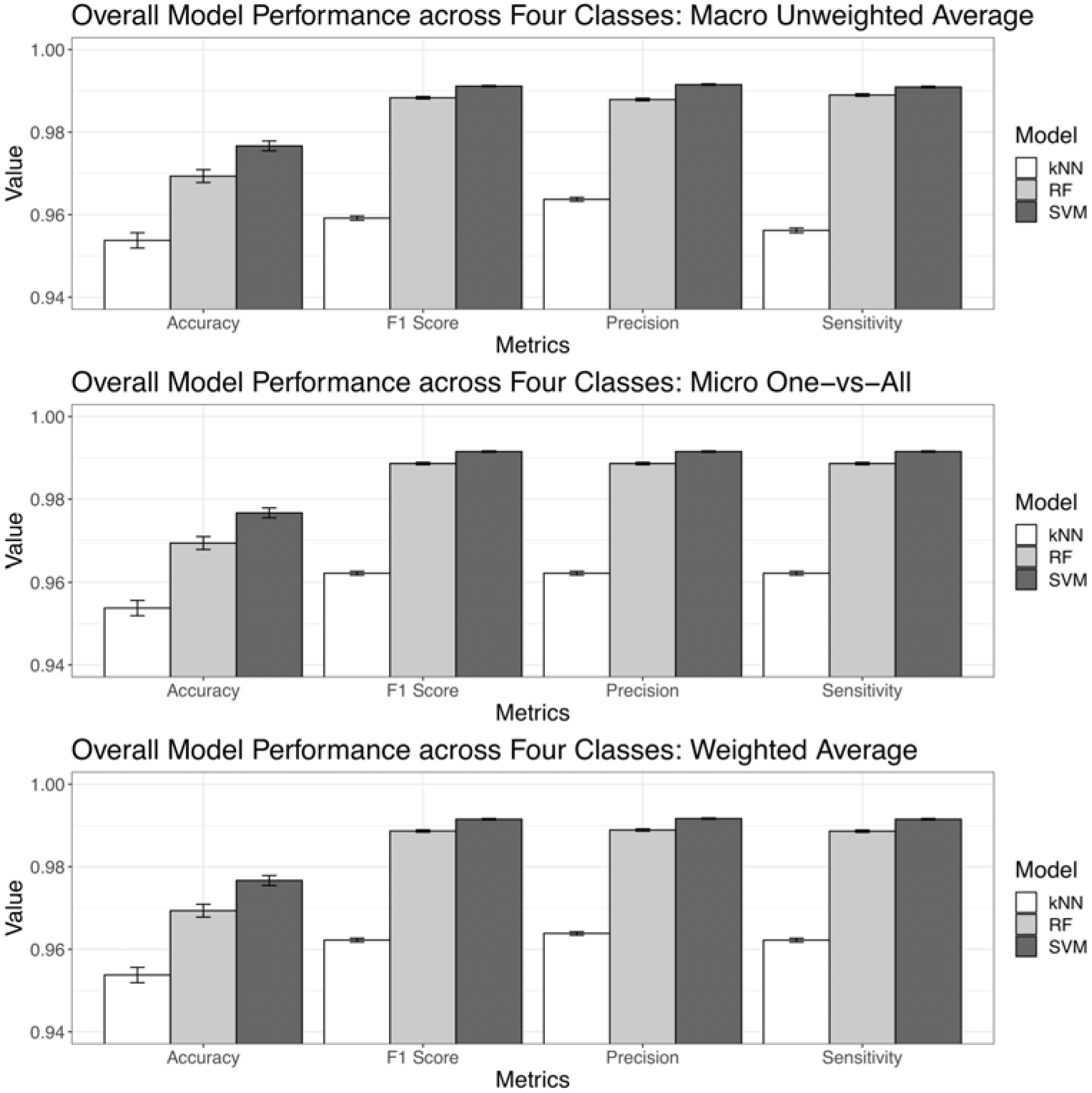
Overall Machine Learning Models Performance across Four Classes.

Note: kNN: k nearest neighbor; RF: random forest; SVM: support vector machine. The performances are based on prediction of the unseen 20% testing data, not on the 80% data to develop the model.

After examining the performance metrics across three different types of ML models, SVM had consistently the best performance with regard to all metrics, followed by RF though the difference was almost indistinguishable. kNN had about 1%-3% deficiency in performance than the other two. We should point out that kNN also had at least 95% accuracy, F1 score, sensitivity, and precision. Therefore, kNN was only bested by two even more competitive models. Furthermore, the relatively small standard errors (SE) demonstrated the robustness of the ML models against different randomly sampled inputs (table S3).

Breaking down to each individual class, healthy class (H) had 95.2%-99.9% prediction accuracy, 95.5%-98.4% F1 score, 91.4%-97.3% sensitivity, and 97.5%-99.9% precision on the testing set depending on specific ML models used (note these were range and not standard errors, which were demonstrated in Fig. 6). The approach to compute class-specific model performance was one-vs-all. For non-severe COVID-19 (NS) class, ML models achieved 95.8%-97.4% accuracy, 97.8%-98.6% F1 score, 99.8% –99.9% sensitivity, and 95.8%-97.4% precision. For severe COVID-19 (S) class, it had 92.4% –99.0% accuracy, 93.4%-96.6% F1 score, 94.3%-94.7% sensitivity, and 92.4%-99.0% precision. For non-COVID viral pneumonia infection (V) class, it had 90.6%-95.0% accuracy, 92.9% –96.8% F1 score, 95.4%-98.8% sensitivity, and 90.6%-95.0% precision. Non-COVID viral infection class (V) was relatively more challenging to differentiate from the other three classes H, NS, and S, but again the difference was not substantial and still justified potential clinical use. Similar to the results in the overall model performance (Fig. 5), class-specific performance metrics also had relatively small SEs, indicating robustness and consistency against randomly selected inputs for training the model. Except for a few classes and model performance metrics, SVM had slightly better performance over RF and kNN. Complete class-specific results were demonstrated in Fig. 6. Complete class-specific performance metrics across three ML models were recorded in supplementary table S4.

**Fig. 6.**
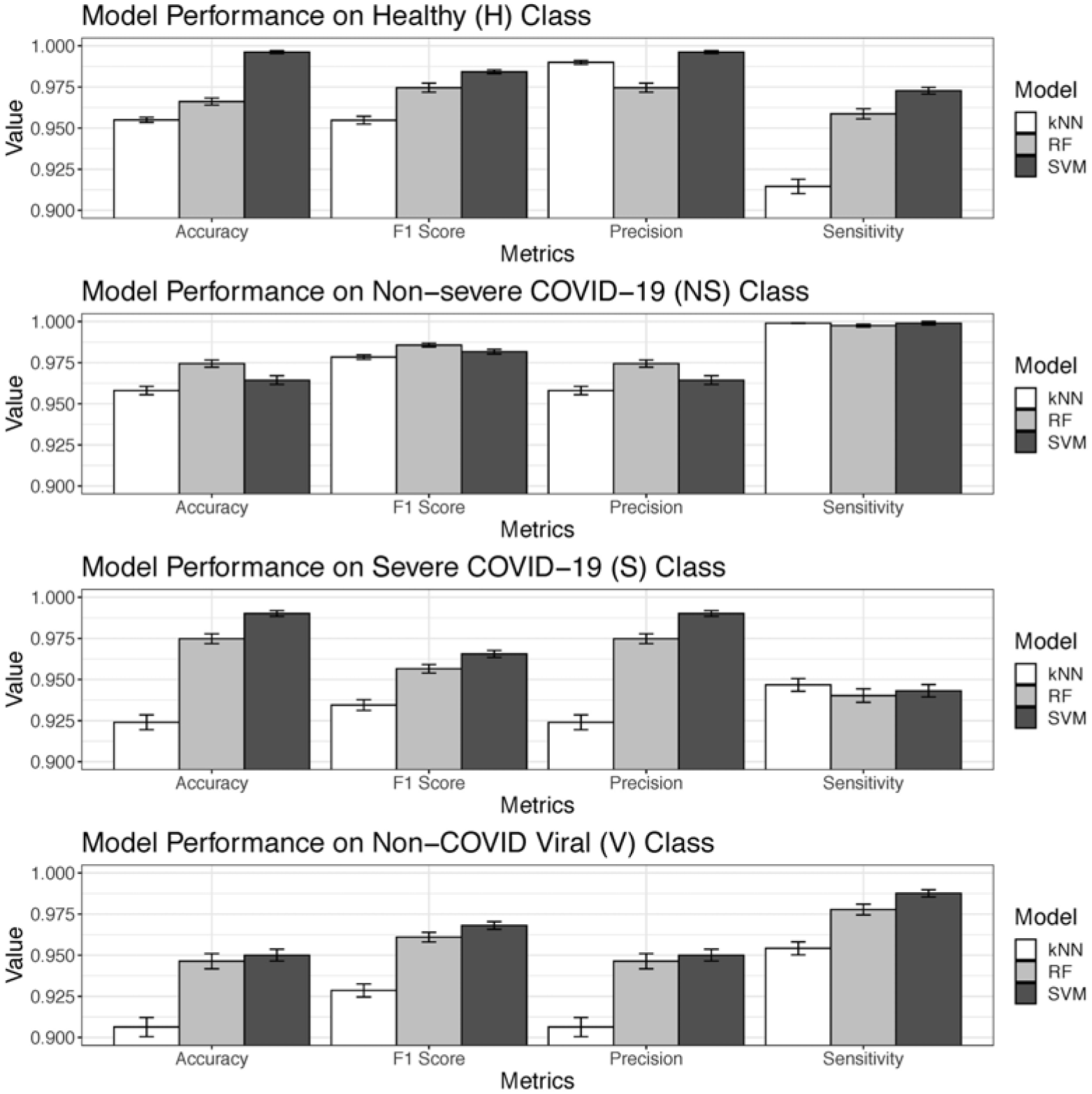
Class-specific Machine Learning Models Performance.

All three ML multinomial classification models based on different computational techniques had consistently high performance both overall (Fig. 5, table S3) and down to specific class (Fig. 6, table S4). Among three types of ML models developed and evaluated, SVM was marginally better than RF and kNN. As a result, ML multinomial classification models were able to accurately differentiate the four classes all at once, provided accurate and more detailed class-specific predictions, and could be reliable decision-making tools for clinical diagnostic support and triaging of suspected COVID-19 patients, who might be non-infected or infected with clinically similar types of virus other than SARS-CoV-2.

In addition to the multimodal classification that incorporated all three different feature sets (binary clinical, continuous lab testing, and CT via CNN; Fig. 1), we also tested how each specific feature modality performed without feature fusion (i.e., unimodality). Using 23 symptom features alone, RF, kNN, and SVM achieved an average of 74.5% (±0.3%), 73.3% (±0.3%), and 75.5% (±0.3%) accuracy on the testing set, respectively. With 10 lab testing features only, RF, kNN, and SVM achieved an average of 67.7% (±0.4%), 56.2% (±0.4%), and 59.5% (±0.3%) accuracy on the testing set, respectively.

The overall accuracy of CNN on CT scan data alone was 90.8% (±0.3%) across the four classes. Further breaking down to each pair of classes, CNN was able to accurately differentiate between severe COVID-19 (S) and healthy (H) classes with 99.9% (<±0.1%) accuracy, between non-COVID viral infection (V) and H with 99.2% (±0.1%) accuracy, between S and NS with 95.4% (±0.1%) accuracy, between non-severe COVID-19 (NS) and healthy (H) with 90.3% (±0.2%) accuracy. However, using CT alone without feature late fusion, it could only differentiate between V and NS with 84.9% (±0.2%) accuracy, and between V and S with 74.2% (±0.2%) accuracy on the testing set.

There were substantial performance boosts by combining input features from different feature modalities and performing multimodal classification than using single feature modality alone. A 15%-42% increase in prediction accuracy on the testing set was achieved compared to single modality models. Note that RF, SVM, and kNN were technically distinct ML models. However, the performance differences among these three distinct ML models were marginal based on multimodal features. Therefore, we concluded that the high performance of COVID-19 classification in this study (Fig. 5 and Fig. 6) was in large part due to multimodal feature late fusion and not by certain types of ML model.

Specifically, Gini impurity scores derived from the RF model had identified major contributing factors that differentiate the four classes. For clinical feature modality, the top three most influential features were having fever, coughing, and elerly (>50yr). The forest plot of odds ratios were provided in Fig. 2 showing the exact influence of these features across classes. For lab testing features, the top three most influential ones were high sensitivity C-reactive protein (hs-CRP), hemoglobin, and absolute count of neutrophil in descending order. The distributions of these three features across four classes as well as multiple comparison results were shown in Fig. 3. While hs-CRP had been a known factor for COVID-19 severity and prognosis (Wang et al. 2020), we further showed that it could also differentiate COVID-19 patients from non-COVID viral pneumonia patients as well as healthy individuals. In addition, different levels of hemoglobin and neutrophils were novel features to accurately distinguish among COVID-19 clinical states, other non-COVID viral pneumonia, and healthy individuals. These results shed light on which set of clinical and lab testing features were the most critical to identify COVID-19 and help guide clinical practice. For CT features extracted from CNN, RF models identified the top three influential ones, namely 6th, 10th, and 1st CT features of the 10-element fully connected layer (Fig. 4). Although these CT features’ actual clinical interpretation was not clear at this time, they showed the promise of accurately differentiating multinomial classes via CT scan all at once, instead of training several CNNs for binary classifications. Future research might reveal more interpretable clinical relevance of these features with COVID-19 pathology.

In summary, different biomedical information across modalities, such as clinical information, lab testing results, and CT scans work synergistically to reveal the multifaceted nature of COVID-19 pathology. Machine learning and deep learning models provided a feasible technical solution of working directly with multimodal biomedical data, and differentiating severe COVID-19, non-severe COVID-19, non-COVID viral infection, and non-infected individuals at the same time with > 97% accuracy.

## Discussion

In this study, we provided a more holistic perspective to characterize COVID-19 and accurately differentiate it (especially non-severe COVID-19 clinical state) from other clinically similar viral pneumonia and non-infections. The human body is an integrated and systemic entity. When infected by pathogens, clinical consequences could be detected not only from biomedical imaging (e.g., CT scans) but also other features such as lab testing results of blood biochemistry (Ferrari et al. 2020, Brinati et al. 2020). One single feature modality might not reveal the entire clinical consequence and provide the best predictive power for COVID-19 detection and classification, but their synergy exceeds the power of any single modality. Currently, multimodality medical data can be effectively stored, transferred, and exchanged in electronic health record systems. The economic cost of acquiring clinical and lab testing modality data were lower than the current confirmatory qRT-PCR. The availability and readiness of these modality would also be an advantage over qRT-PCR which has a long turnaround time right now. This study unleashed the power of multimodality medical information for an emerging pandemic where confirmatory molecular tests had reliability and availability issues across time and space. This novel application could be utilized in disease preparedness of incoming waves of epidemics in the future.

Upon further examining the comprehensive patient symptom data, we suggested that our current understanding and definition of “asymptomatic” COVID-19 would be inadequate. Among 214 non-severe COVID-19 patients, 60 had no fever (< 37C), 78 did not cough, 141 did not feel chest congestion and pain, and 172 reported no sore throat upon admission. There were 10 confirmed patients (5% of non-severe class) presenting none of these common symptoms and could be considered as “asymptomatic” COVID-19 patients. Even further considering headache, muscle pain, and fatigue, there were still 4 patients (2%) showing no symptoms related to typical respiratory diseases. One of these four patients, however, had diarrhea upon admission. Therefore, using symptom features alone would not be sufficient to detect and differentiate these asymptomatic COVID-19 patients. Nevertheless, all these asymptomatic patients had been successfully detected via our model with no false negatives. This finding showed the incompleteness of the current definition and understanding of “asymptomatic” COVID-19, and the potential power of non-traditional analytical tools to identify these patients.

Following this perspective, we further developed a comprehensive end-to-end analytical framework that integrated both high-dimensional biomedical imaging and low-dimensional clinical and lab testing data. CT scans were first processed by deep learning convolutional neural networks (CNN). We developed a customized ResNet CNN architecture with two fully connected (FC) layers before the final output layer. We then used the 2nd FC layer as the low-dimensional representation of the original high-dimensional CT data. In other words, CNN was applied first for dimensional reduction. Feature fusion of CT (represented by FC layer), clinical, and lab testing feature modalities demonstrated feasibility and high accuracy in differentiating non-severe, severe COVID-19, non-COVID viral pneumonia, and healthy clases all at once. The consistent high performance across three different types of ML models, RF, SVM, and kNN, as well as substantial performance boost from using single modality further unleashed the hidden power of feature fusion across different biomedical feature modalities. Feature fusion of multimodal biomedical data significantly boosted prediction accuracy (>97%) on testing set from using any single feature modality alone (60%-80%).

We compared our model performance based on the multimodal biomedical data of 683 participants against state-of-the-art benchmarks of COVID-19 classification studies. DL studies with thoracic CT based on 87 participants claimed to have > 99% accuracy (Ko et al. 2020) and 86%-99% accuracy on 200 participants (Ardakani et al. 2020) between COVID-19 patients and non-COVID individuals. Another study recorded 95% AUC to differentiate COVID-19 and other community-acquired pneumonia (CAP) in 3322 participants (Li et al. 2020). 92% area under curve (AUC) was achieved in a study with 905 COVID-19 and non-COVID participants utilizing multimodal CT, clinical information, and lab testing information (Mei et al. 2020). A study using CT scans to differentiate three multinomial classes: COVID (no clinical state information), non-COVID viral pneumonia, and health individuals had 89%-96% accuracy based on a total of 230 participants (Hu et al. 2020). In addition, professionally trained human radiologists could achieve 60%-83% accuracy on differentiating COVID-19 from other types of CAP (Bai et al. 2020). Therefore, our model performance would be on par or even better than these benchmark models and exceed the performance of human radiologist. Moreover, existing studies generally focused on differentiating COVID-19 patients from non-COVID individuals or other types of pneumonia patients. In other words, current COVID-19 classification models were mostly binary classifiers. Our study not only detected COVID-19 from healthy individuals but answered the more important clinical question to differentiate COVID-19 from other viral infections, as well as distinguish different clinical states (severe vs non-severe) of COVID-19. Therefore, it was a novel and effective breakthrough for clinical applications, not just incremental improvement over existing ML models.

Success of this study sheds light on many other disease systems that have multimodal biomedical data input. Specifically, feature fusion between high- and low-dimensional biomedical data modalities can be applied to more feature modalities, for example, individual-level high-dimensional “-omics” data. Currently, genome-wide association study (GWAS) between individual single nucleotide polymorphism (SNP) and COVID-19 susceptibility has revealed several target loci that are involved in COVID-19 pathology. Following a similar approach, we may also perform dimensional reduction of “-omics” data first, and then perform data fusion with other low-dimensional modalities (Tarnok 2020, Ray and Srivastava 2020, Arga 2020).

The classification part of this study adopts a hybrid of deep learning (CNN) and machine learning (RF, SVM, kNN) models via feature late fusion. By using various data-driven methods, it avoids the potential pitfall of cause-effect and focuses directly on the more important clinical question. For instance, many comorbidities such as diabetes (Wicaksana et al. 2020, Abdi et al. 2020) and cardiovascular diseases (Madjid et al. 2020, Matsushita et al. 2020) are strongly associated with occurrence of severe COVID-19. It is still unclear whether diabetes (or reduced kidney function) causes severe COVID-19, or COVID-19 infection worsens existing diabetes, or they are actually mutually influential to each other and result in undesirable clinical prognosis. Future studies can use data-driven methods to further investigate the causality of comorbidities and COVID-19.

There are some limitations of this study. First, in order to perform *multinomial* classification across the four classes, we have to discard a lot of features especially in lab testing modality. The non-COVID viral pneumonia (V) class used a different electronic health record (EHR) system that collected different lab testing features from participants in Wuhan (COVID and healthy classes). Many lab testing features were demonstrated to accurately differentiate severe and non-severe COVID-19 in our previous investigation, such as hsTNI, ddimer, LDH, etc. However these features were not present (or largely missing) in the V class. Eventually, only 10 lab testing features were included, compared to an average of 20–30 features available in different EHR systems. This is probably the reason why lab testing feature modality alone was not able to make accurate classification (highest accuracy 67.7% using RF) across all four classes in this specific study. In addition, although we had a reasonably large participant pool of 638 individuals, more participants were expected to further validate the findings of this study.

Another potential pitfall is that not all feature modalities could be readily available at the same time for feature fusion and multimodal classification. For single modality features, CT is the best performer to generate accurate predictions. However, CT is usually performed in the radiology department. Lab testing may be outsourced and also takes time for the results to turn in. Consequently, there might be lags of data availability among different feature modalities. We suggest that when multimodal features are not available all at once, single modality features can be used to make first-round triaging. Multimodal features are needed when accuracy is a must.

Finally, although the participants in this study came from different healthcare facilities, the majority of them were still Chinese Han ethnicity. The biomedical features among different COVID-19 and non-COVID classes may be different in other races, ethnicity, or other confounding factors. Cross-validation of the findings in this study based on other ethnicity groups and larger sample size is needed for future research.

## Data Availability

All codes and de-identified data files were freely available on GitHub.

## Acknowledgments

This study is supported by the North Carolina Biotechnology Center Flash Grant on COVID-19 Clinical Research (2020-FLG-3898), the National Science Foundation for Young Scientists of China (81703201), the Natural Science Foundation for Young Scientists of Jiangsu Province (BK20171076), the Jiangsu Provincial Medical Innovation Team (CXTDA2017029), the Jiangsu Provincial Medical Youth Talent program (QNRC2016548), the Jiangsu Preventive Medicine Association program (Y2018086), the Lifting Program of Jiangsu Provincial Scientific and Technological Association, and the Jiangsu Government Scholarship for Overseas Studies.

## Author contributions

M.X. designed the study. L.O. and J.L. derived and processed data. L.O., F.S.B., Q.L., L.H., B.Z, J.L., S.C. interpreted results. L.S., F.S.B. S.C. performed analyses. Y.G. and S.C. supervised. S.C. developed the manuscript.

---

**Table S1.**
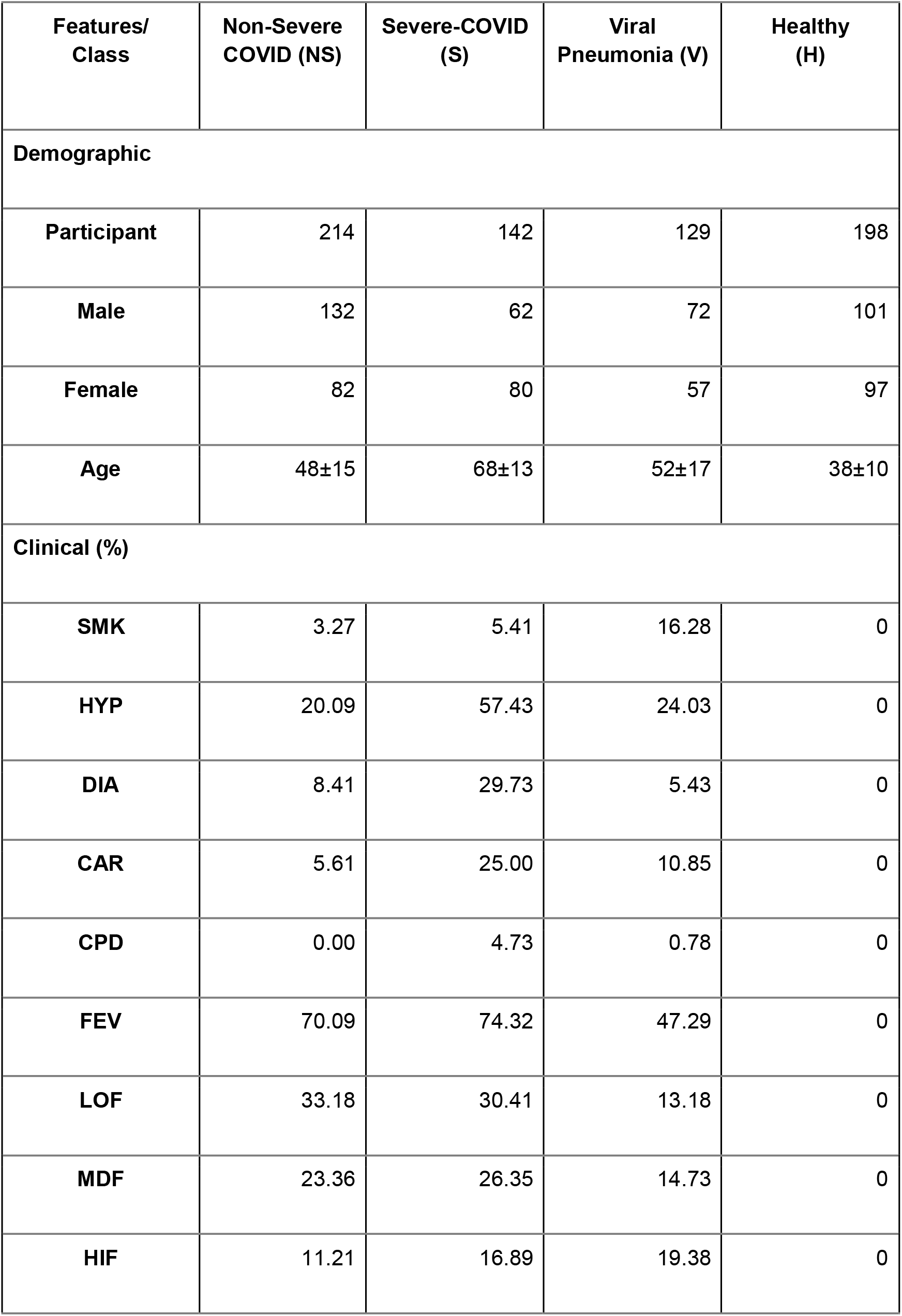
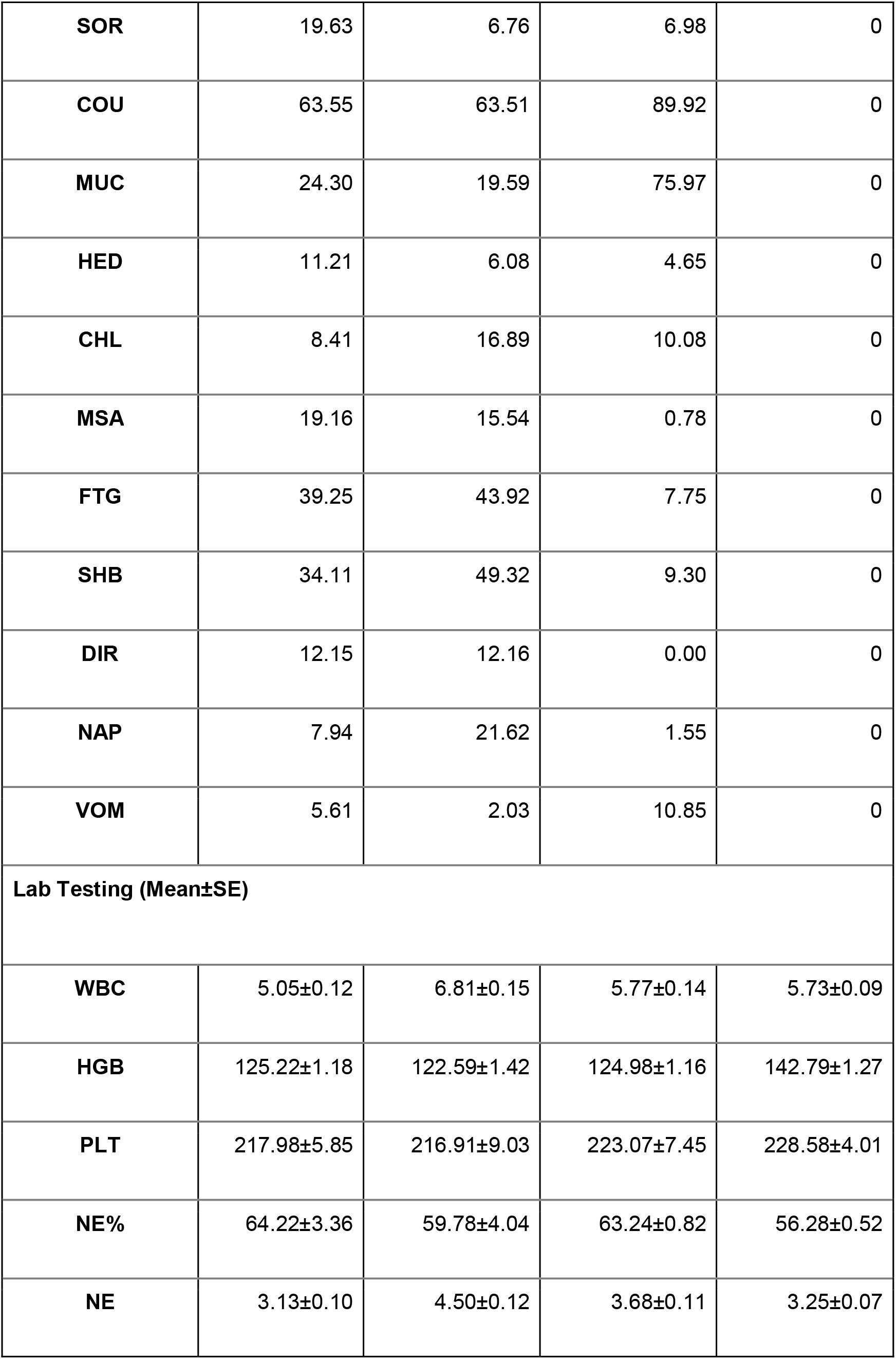
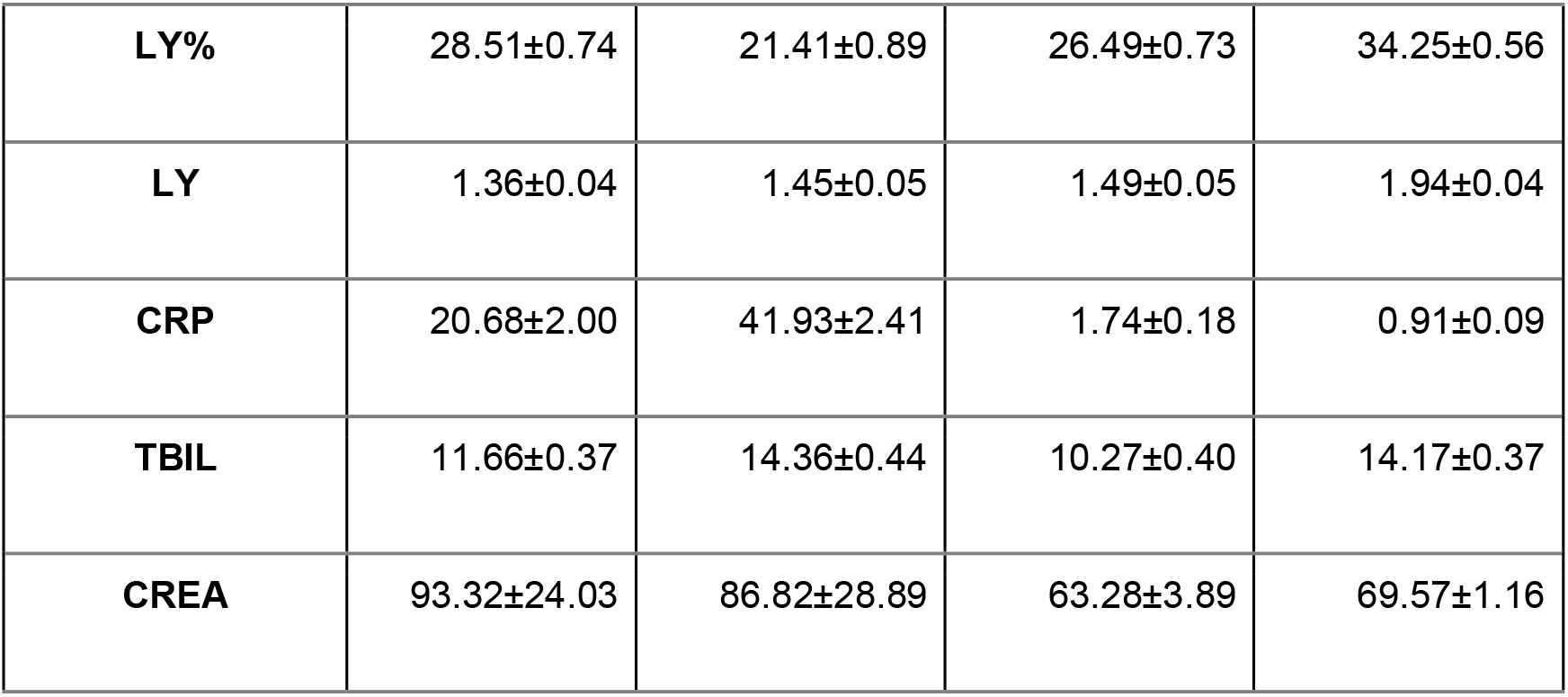
Demographic, Clinical, and Lab Testing Results among Four Classes.

| **Features/**<br>**Class** | **Non-Severe**<br>**COVID (NS)** | **Severe-COVID**<br>**(S)** | **Viral**<br>**Pneumonia (V)** | **Healthy**<br>**(H)** |
| --- | --- | --- | --- | --- |
| **Demographic** |  |  |  |  |
| **Participant** | 214 | 142 | 129 | 198 |
| **Male** | 132 | 62 | 72 | 101 |
| **Female** | 82 | 80 | 57 | 97 |
| **Age** | 48±15 | 68±13 | 52±17 | 38±10 |
| **Clinical (%)** |  |  |  |  |
| **SMK** | 3.27 | 5.41 | 16.28 | 0 |
| **HYP** | 20.09 | 57.43 | 24.03 | 0 |
| **DIA** | 8.41 | 29.73 | 5.43 | 0 |
| **CAR** | 5.61 | 25.00 | 10.85 | 0 |
| **CPD** | 0.00 | 4.73 | 0.78 | 0 |
| **FEV** | 70.09 | 74.32 | 47.29 | 0 |
| **LOF** | 33.18 | 30.41 | 13.18 | 0 |
| **MDF** | 23.36 | 26.35 | 14.73 | 0 |
| **HIF** | 11.21 | 16.89 | 19.38 | 0 |
| **SOR** | 19.63 | 6.76 | 6.98 | 0 |
| **COU** | 63.55 | 63.51 | 89.92 | 0 |
| **MUC** | 24.30 | 19.59 | 75.97 | 0 |
| **HED** | 11.21 | 6.08 | 4.65 | 0 |
| **CHL** | 8.41 | 16.89 | 10.08 | 0 |
| **MSA** | 19.16 | 15.54 | 0.78 | 0 |
| **FTG** | 39.25 | 43.92 | 7.75 | 0 |
| **SHB** | 34.11 | 49.32 | 9.30 | 0 |
| **DIR** | 12.15 | 12.16 | 0.00 | 0 |
| **NAP** | 7.94 | 21.62 | 1.55 | 0 |
| **VOM** | 5.61 | 2.03 | 10.85 | 0 |
| **Lab Testing (Mean±SE)** |  |  |  |  |
| **WBC** | 5.05±0.12 | 6.81±0.15 | 5.77±0.14 | 5.73±0.09 |
| **HGB** | 125.22±1.18 | 122.59±1.42 | 124.98±1.16 | 142.79±1.27 |
| **PLT** | 217.98±5.85 | 216.91±9.03 | 223.07±7.45 | 228.58±4.01 |
| **NE%** | 64.22±3.36 | 59.78±4.04 | 63.24±0.82 | 56.28±0.52 |
| **NE** | 3.13±0.10 | 4.50±0.12 | 3.68±0.11 | 3.25±0.07 |
| **LY%** | 28.51±0.74 | 21.41±0.89 | 26.49±0.73 | 34.25±0.56 |
| **LY** | 1.36±0.04 | 1.45±0.05 | 1.49±0.05 | 1.94±0.04 |
| **CRP** | 20.68±2.00 | 41.93±2.41 | 1.74±0.18 | 0.91±0.09 |
| **TBIL** | 11.66±0.37 | 14.36±0.44 | 10.27±0.40 | 14.17±0.37 |
| **CREA** | 93.32±24.03 | 86.82±28.89 | 63.28±3.89 | 69.57±1.16 |

Note: age (dichotomized with threshold of 50yr) and gender were used as clinical features in subsequent analyses.

**Table S2.**
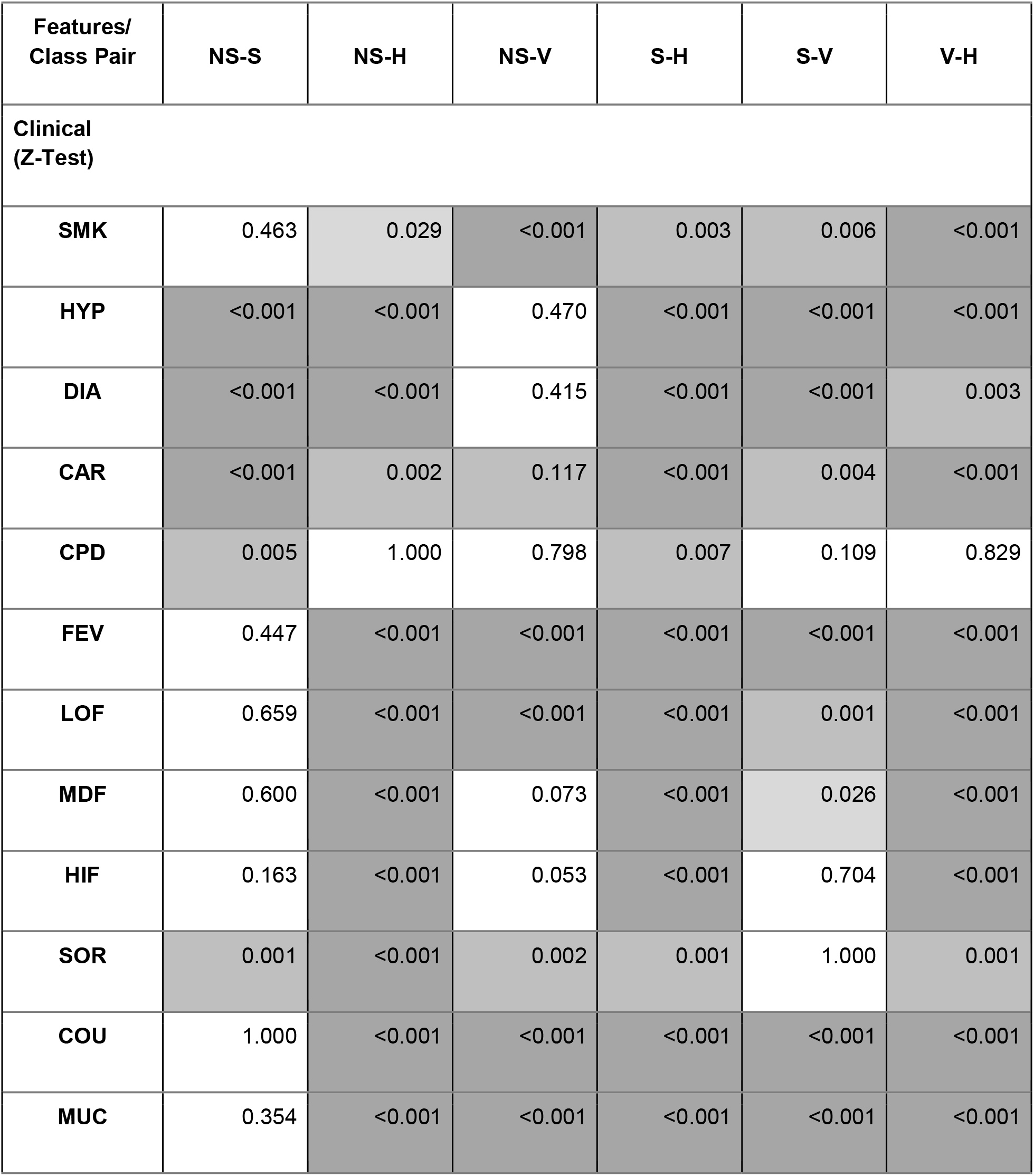
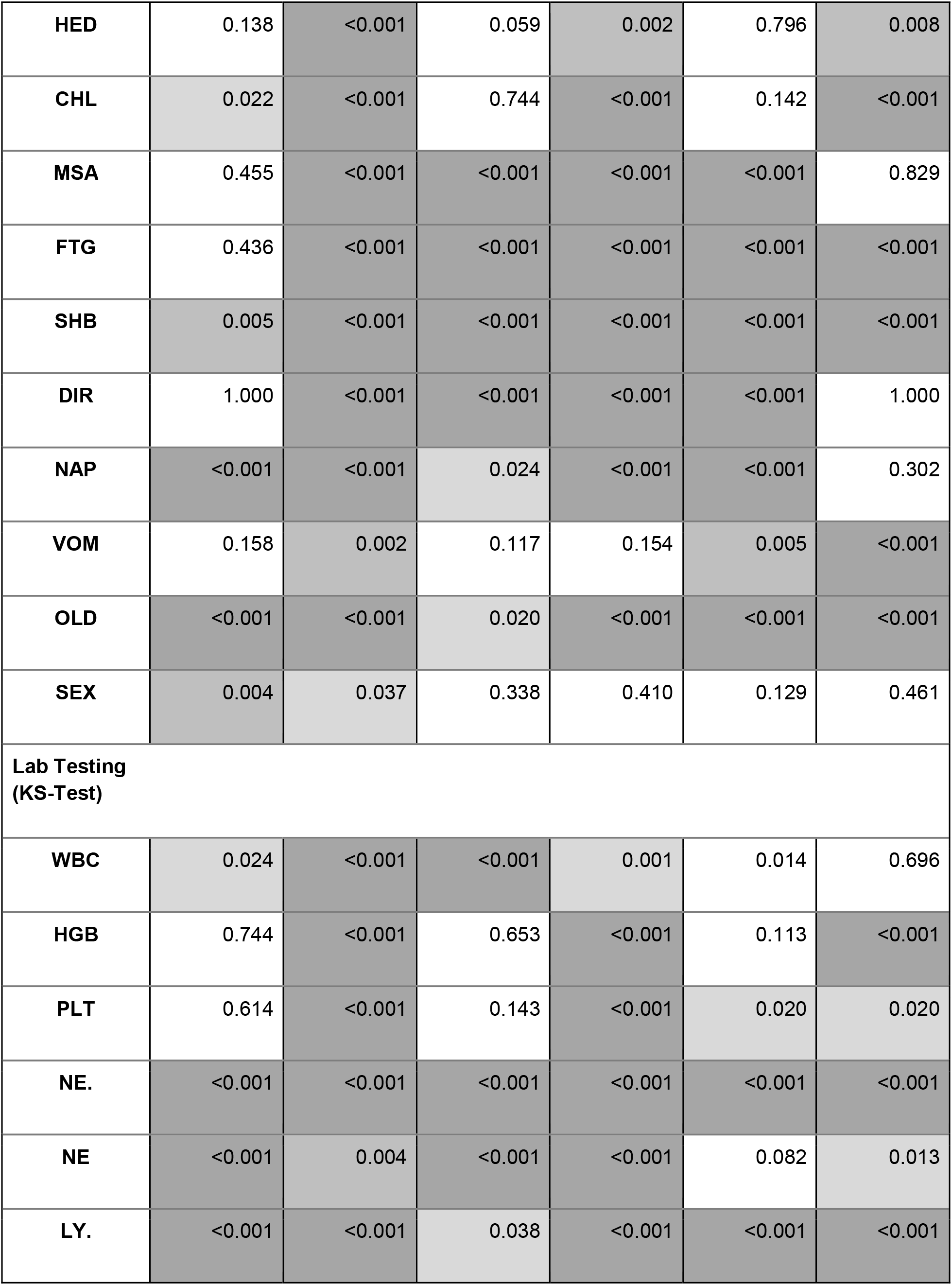
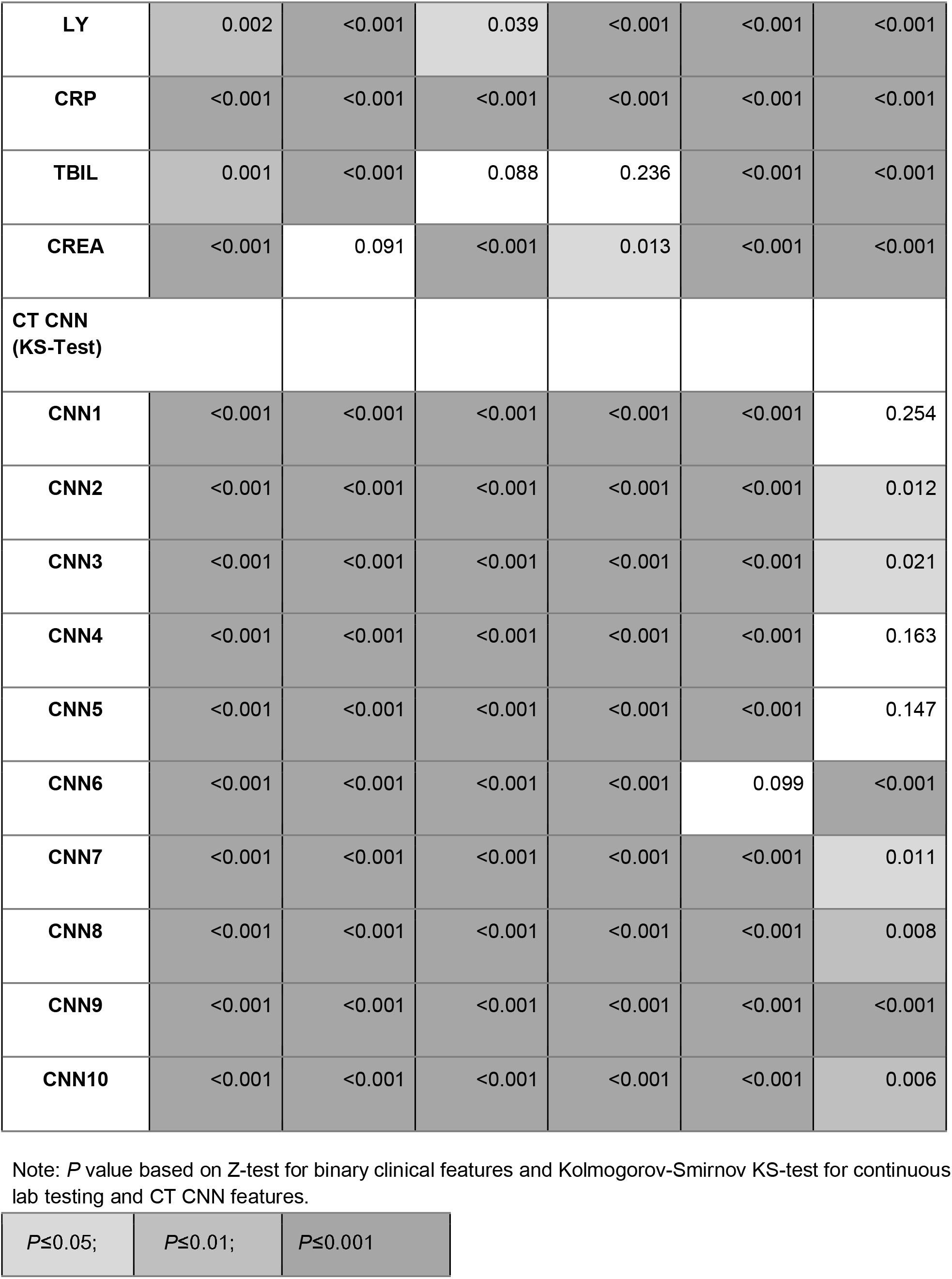
Z-Test and/or Kolmogorov-Smirnov Test Results of Significance for Each Feature among Four Classes.

| **Features/**<br>**Class Pair** | **NS-S** | **NS-H** | **NS-V** | **S-H** | **S-V** | **V-H** |
| --- | --- | --- | --- | --- | --- | --- |
| **Clinical**<br>**(Z-Test)** |  |  |  |  |  |  |
| **SMK** | 0.463 | 0.029 | <0.001 | 0.003 | 0.006 | <0.001 |
| **HYP** | <0.001 | <0.001 | 0.470 | <0.001 | <0.001 | <0.001 |
| **DIA** | <0.001 | <0.001 | 0.415 | <0.001 | <0.001 | 0.003 |
| **CAR** | <0.001 | 0.002 | 0.117 | <0.001 | 0.004 | <0.001 |
| **CPD** | 0.005 | 1.000 | 0.798 | 0.007 | 0.109 | 0.829 |
| **FEV** | 0.447 | <0.001 | <0.001 | <0.001 | <0.001 | <0.001 |
| **LOF** | 0.659 | <0.001 | <0.001 | <0.001 | 0.001 | <0.001 |
| **MDF** | 0.600 | <0.001 | 0.073 | <0.001 | 0.026 | <0.001 |
| **HIF** | 0.163 | <0.001 | 0.053 | <0.001 | 0.704 | <0.001 |
| **SOR** | 0.001 | <0.001 | 0.002 | 0.001 | 1.000 | 0.001 |
| **COU** | 1.000 | <0.001 | <0.001 | <0.001 | <0.001 | <0.001 |
| **MUC** | 0.354 | <0.001 | <0.001 | <0.001 | <0.001 | <0.001 |
| **HED** | 0.138 | <0.001 | 0.059 | 0.002 | 0.796 | 0.008 |
| **CHL** | 0.022 | <0.001 | 0.744 | <0.001 | 0.142 | <0.001 |
| **MSA** | 0.455 | <0.001 | <0.001 | <0.001 | <0.001 | 0.829 |
| **FTG** | 0.436 | <0.001 | <0.001 | <0.001 | <0.001 | <0.001 |
| **SHB** | 0.005 | <0.001 | <0.001 | <0.001 | <0.001 | <0.001 |
| **DIR** | 1.000 | <0.001 | <0.001 | <0.001 | <0.001 | 1.000 |
| **NAP** | <0.001 | <0.001 | 0.024 | <0.001 | <0.001 | 0.302 |
| **VOM** | 0.158 | 0.002 | 0.117 | 0.154 | 0.005 | <0.001 |
| **OLD** | <0.001 | <0.001 | 0.020 | <0.001 | <0.001 | <0.001 |
| **SEX** | 0.004 | 0.037 | 0.338 | 0.410 | 0.129 | 0.461 |
| **Lab Testing**<br>**(KS-Test)** |  |  |  |  |  |  |
| **WBC** | 0.024 | <0.001 | <0.001 | 0.001 | 0.014 | 0.696 |
| **HGB** | 0.744 | <0.001 | 0.653 | <0.001 | 0.113 | <0.001 |
| **PLT** | 0.614 | <0.001 | 0.143 | <0.001 | 0.020 | 0.020 |
| **NE.** | <0.001 | <0.001 | <0.001 | <0.001 | <0.001 | <0.001 |
| **NE** | <0.001 | 0.004 | <0.001 | <0.001 | 0.082 | 0.013 |
| **LY.** | <0.001 | <0.001 | 0.038 | <0.001 | <0.001 | <0.001 |
| **LY** | 0.002 | <0.001 | 0.039 | <0.001 | <0.001 | <0.001 |
| **CRP** | <0.001 | <0.001 | <0.001 | <0.001 | <0.001 | <0.001 |
| **TBIL** | 0.001 | <0.001 | 0.088 | 0.236 | <0.001 | <0.001 |
| **CREA** | <0.001 | 0.091 | <0.001 | 0.013 | <0.001 | <0.001 |
| **CT CNN**<br>**(KS-Test)** |  |  |  |  |  |  |
| **CNN1** | <0.001 | <0.001 | <0.001 | <0.001 | <0.001 | 0.254 |
| **CNN2** | <0.001 | <0.001 | <0.001 | <0.001 | <0.001 | 0.012 |
| **CNN3** | <0.001 | <0.001 | <0.001 | <0.001 | <0.001 | 0.021 |
| **CNN4** | <0.001 | <0.001 | <0.001 | <0.001 | <0.001 | 0.163 |
| **CNN5** | <0.001 | <0.001 | <0.001 | <0.001 | <0.001 | 0.147 |
| **CNN6** | <0.001 | <0.001 | <0.001 | <0.001 | 0.099 | <0.001 |
| **CNN7** | <0.001 | <0.001 | <0.001 | <0.001 | <0.001 | 0.011 |
| **CNN8** | <0.001 | <0.001 | <0.001 | <0.001 | <0.001 | 0.008 |
| **CNN9** | <0.001 | <0.001 | <0.001 | <0.001 | <0.001 | <0.001 |
| **CNN10** | <0.001 | <0.001 | <0.001 | <0.001 | <0.001 | 0.006 |

Note: *P* value based on Z-test for binary clinical features and Kolmogorov-Smirnov KS-test for continuous lab testing and CT CNN features.

|  |  |  |
| --- | --- | --- |
| *P*≤0.05; | *P*≤0.01; | *P*≤0.001 |

**Table S3.**
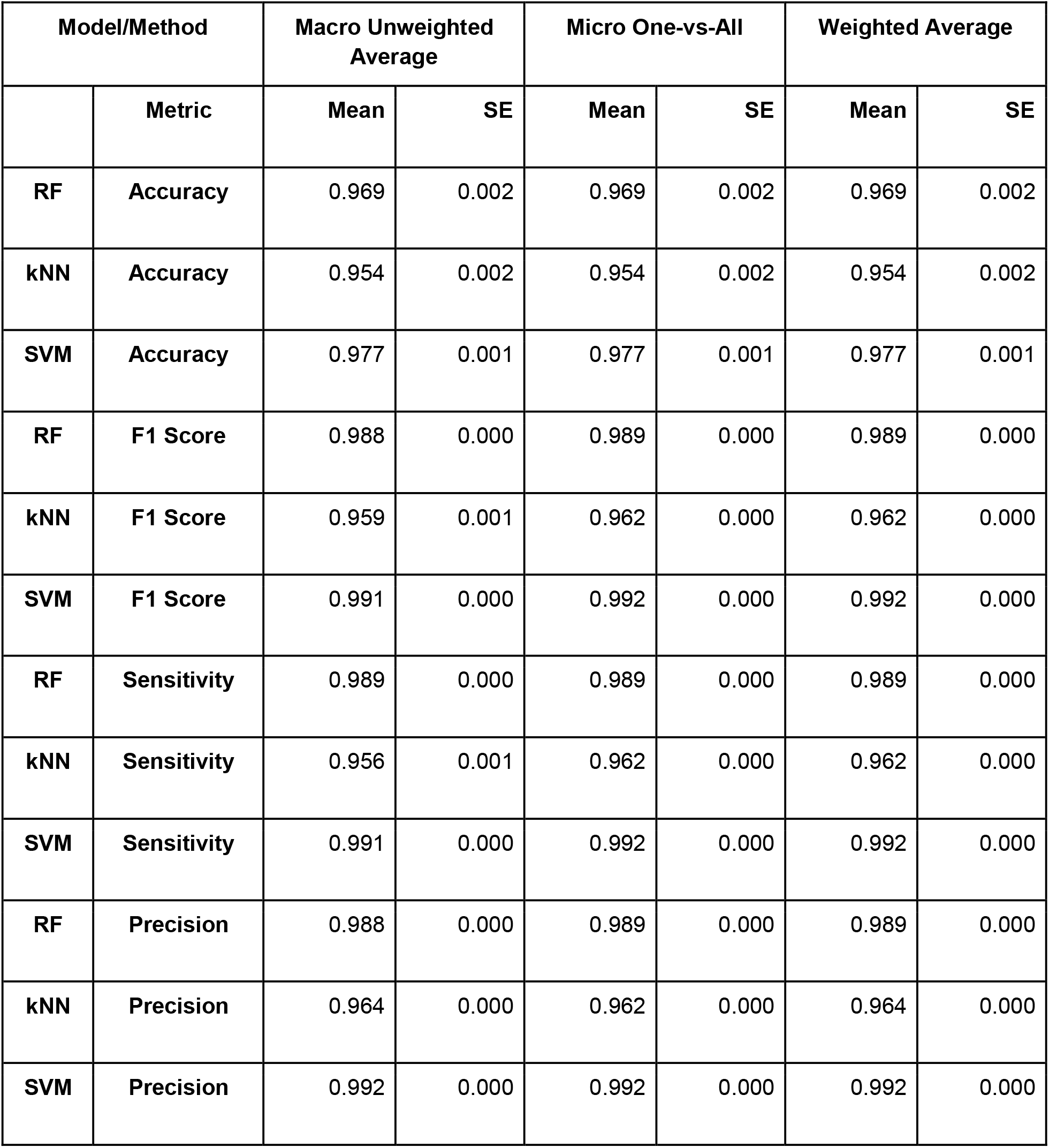
Overall Machine Learning Model Performance Comparison.

**Table S4.** Class-specific Machine Learning Model Performance Comparison.

| Model/Class |  | Severe COVID-19 (S) |  | Non-severe COVID-19 (NS) |  | Non-COVID Viral Pneumonia (V) |  | Non-Infected Healthy (H) |  |
| --- | --- | --- | --- | --- | --- | --- | --- | --- | --- |
|  | Metric | Mean | SE | Mean | SE | Mean | SE | Mean | SE |
| RF | Accuracy | 0.975 | 0.003 | 0.974 | 0.002 | 0.946 | 0.005 | 0.966 | 0.002 |
| kNN | Accuracy | 0.924 | 0.005 | 0.958 | 0.003 | 0.906 | 0.006 | 0.955 | 0.002 |
| SVM | Accuracy | 0.990 | 0.002 | 0.964 | 0.003 | 0.950 | 0.004 | 0.996 | 0.001 |
| RF | F1 Score | 0.957 | 0.003 | 0.986 | 0.001 | 0.961 | 0.003 | 0.975 | 0.003 |
| kNN | F1 Score | 0.934 | 0.003 | 0.978 | 0.001 | 0.929 | 0.004 | 0.955 | 0.002 |
| SVM | F1 Score | 0.966 | 0.002 | 0.982 | 0.001 | 0.968 | 0.002 | 0.984 | 0.001 |
| RF | Sensitivity | 0.940 | 0.004 | 0.998 | 0.001 | 0.978 | 0.003 | 0.959 | 0.003 |
| kNN | Sensitivity | 0.947 | 0.004 | 0.999 | 0.000 | 0.954 | 0.004 | 0.915 | 0.004 |
| SVM | Sensitivity | 0.943 | 0.004 | 0.999 | 0.001 | 0.988 | 0.002 | 0.973 | 0.002 |
| RF | Precision | 0.975 | 0.003 | 0.974 | 0.002 | 0.946 | 0.005 | 0.975 | 0.003 |
| kNN | Precision | 0.924 | 0.005 | 0.958 | 0.003 | 0.906 | 0.006 | 0.990 | 0.001 |
| SVM | Precision | 0.990 | 0.002 | 0.964 | 0.003 | 0.950 | 0.004 | 0.996 | 0.001 |

**Fig. S1.**
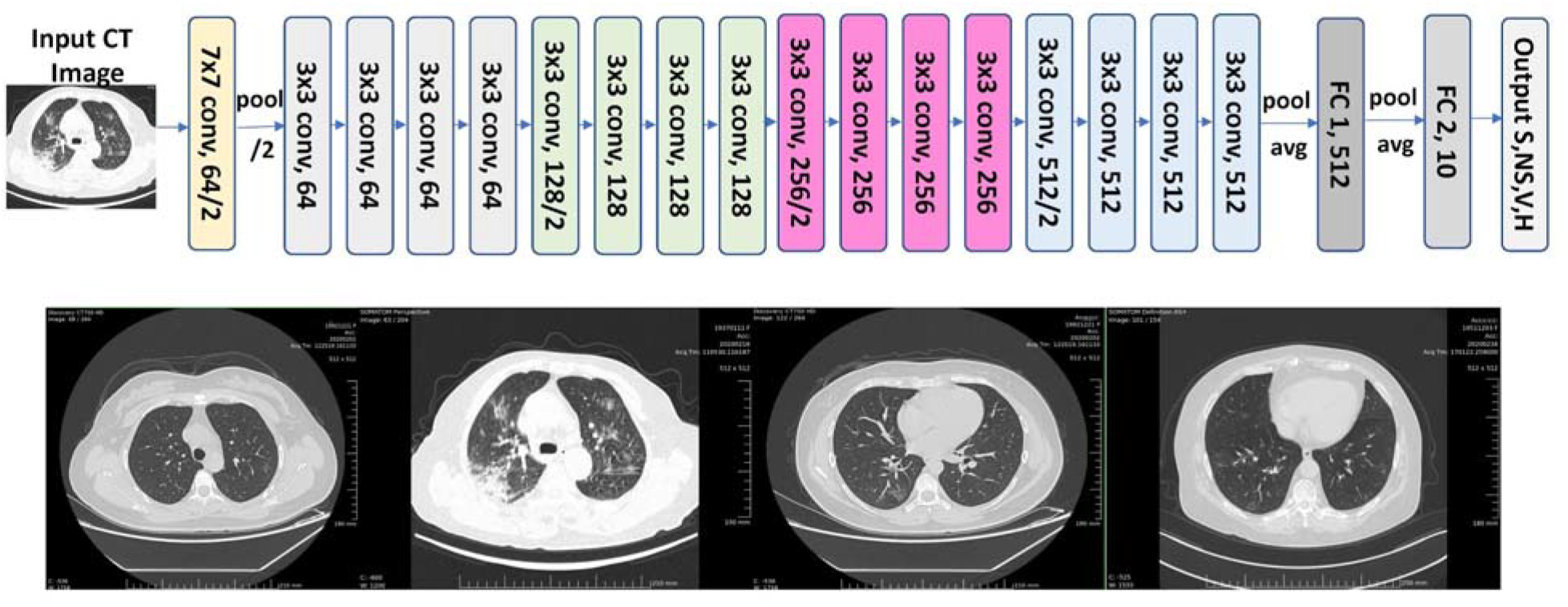
Architecture of Customized ResNet-18 CNN and Sample CT Scans of Four Classes.

Note: left: non-severe COVID-19 (NS); middle-left: severe COVID-19 (S) showing typical ground glass opaque (GGO); middle-right: non-COVID viral pneumonia (V); right: non-infected healthy (H).

**Fig. S2.**
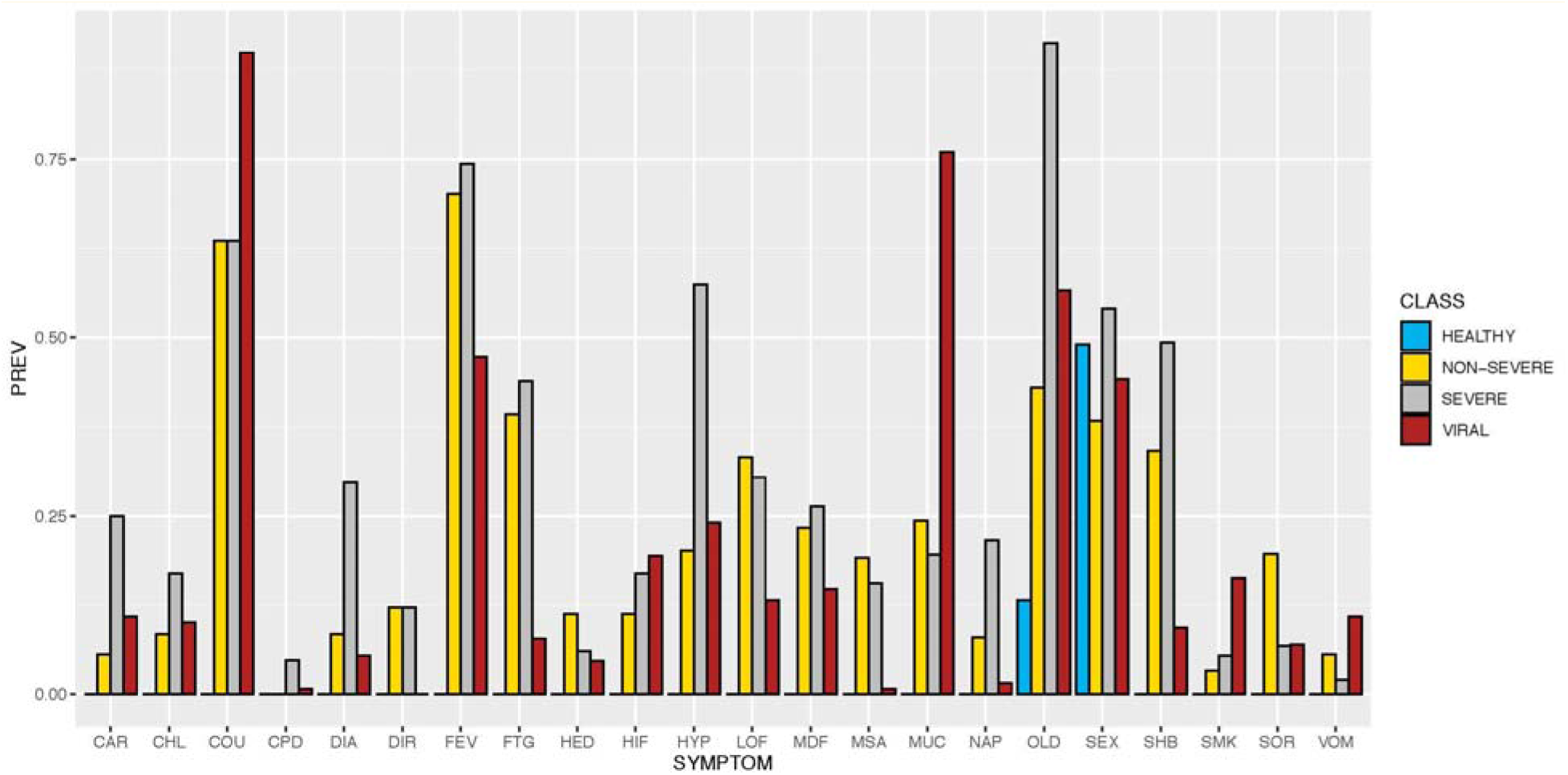
Clinical Features Comparison across Four Classes.

Note: these features had 0–1 binary values so Y-axis was prevalence (percentage) instead of continuous numeric values of mean and standard errors.

**Fig. S3.**
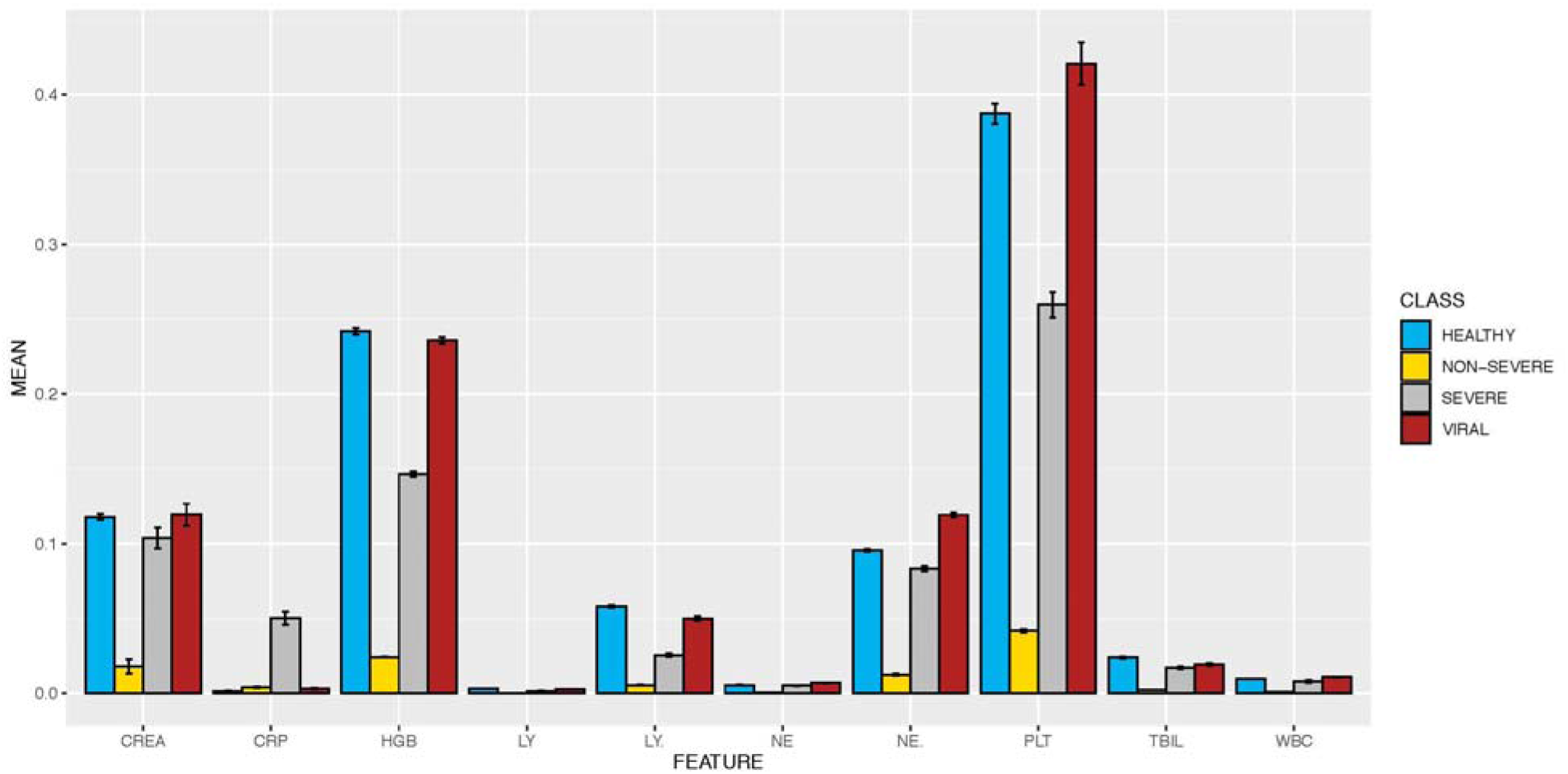
Lab Testing Features Comparison across Four Classes.

**Fig. S4.**
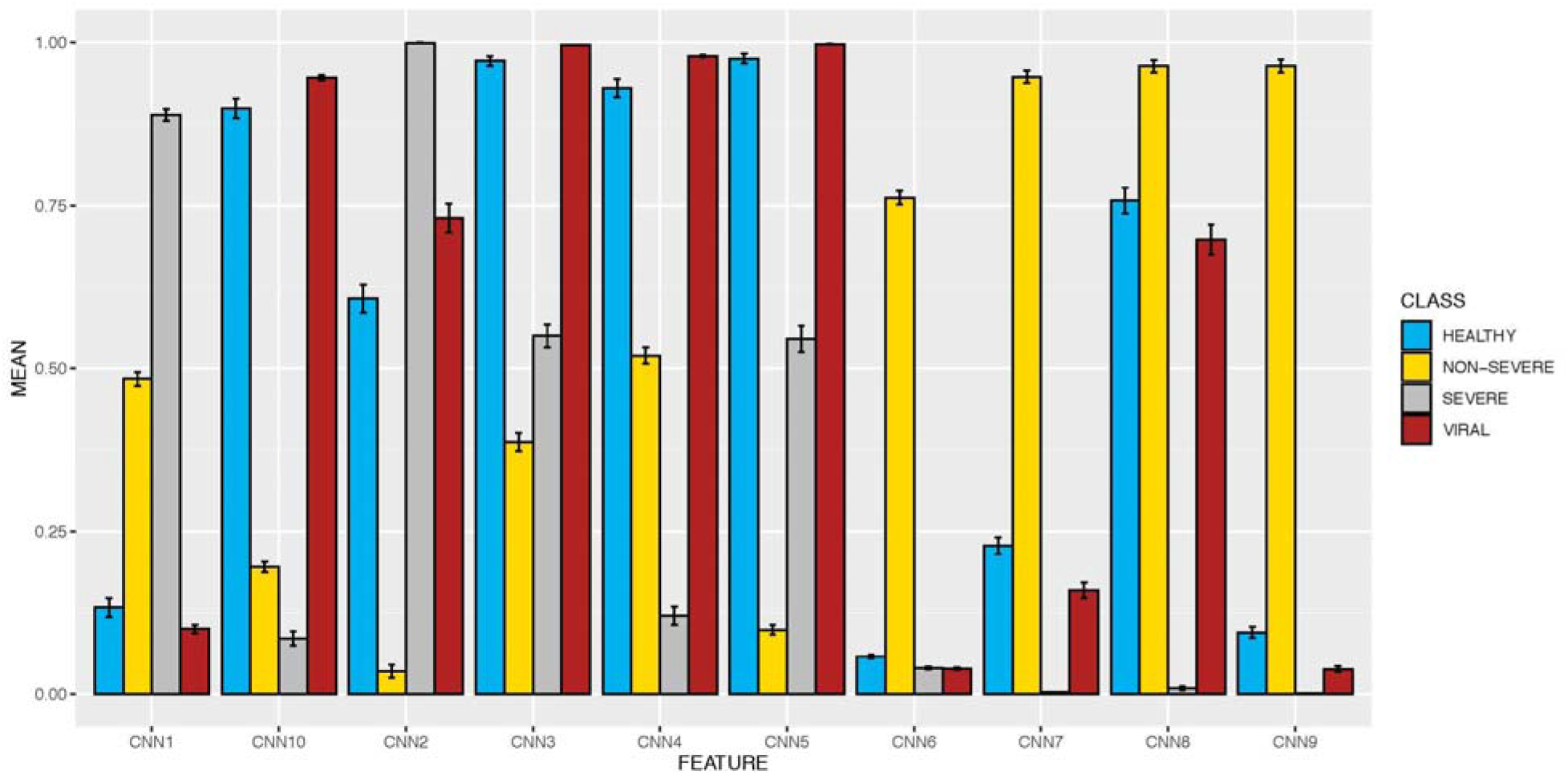
CT Features Extracted via Deep Learning Convolutional Neural Network (CNN) Comparison across Four Classes.

